# Uncovering the Heritable Components of Multimorbidities and Disease Trajectories: A Nationwide Cohort Study

**DOI:** 10.1101/2023.02.08.23285642

**Authors:** David Westergaard, Frederik Hytting Jørgensen, Jens Waaben, Mette Lademann, Thomas Folkmann Hansen, Jolien Cremers, Sisse Rye Ostrowski, Ole Birger Vesterager Pedersen, Danish Blood Donor Study Genomic Consortium, Roc Requant, Isabella Friis Jørgensen, Tom Fitzgerald, Ewan Birney, Karina Banasik, Laust Mortensen, Søren Brunak

## Abstract

Quantifying the contribution of genetics and environmental effects on disease initiation and progression, as well as the shared genetics of different diseases, is vital for the understanding of the disease etiology of multimorbidities. In this study, we leverage nationwide Danish registries to provide a granular atlas of the genetic origin of disease phenotypes for a cohort of all Danes 1978-2018 with partially known pedigree (n = 6.3 million). We estimate the heritability and genetic correlation between thousands of disease phenotypes using a novel approach that can be scaled to nationwide data. Our findings confirm the importance of genetics for a number of known associations and increase the resolution of heritability by adding numerous novel associations, some of which point to shared biologically origin of different phenotypes. We also establish the heritability of disease trajectories and the importance of sex-specific genetic contributions.

## Introduction

Quantifying the contribution of genetics and environmental effects on disease initiation and progression, as well as the shared genetics between diseases, is vital to understanding disease etiology and our ability to customize clinical treatment.^1–4^ The role of shared environment or genetics in the etiology shared between diseases is understudied. This is unfortunate, as shared etiology is likely to drive comorbidity and thus influence disease trajectories.^5^ Small populations, incomplete data, and short follow-up time have, in combination, limited comprehensive research in this life-course perspective. Thus, resolving the contribution of genetics in temporally, comorbid diseases and multimorbid disease trajectories remains a challenge.^6,7^

Heritability is an important concept in partitioning environmental and biological factors with a long history in genetics and medicine.^2^ Several approaches exist for determining heritability. Most commonly used are twin studies, where the phenotype concordance rate is compared between monozygotic and dizygotic twin pairs. However, twin cohorts are of limited availability and size; thus, obtaining sufficiently powered sample sizes for studying late-onset diseases and disease trajectories is difficult. However, the exponential growth and availability of cohorts with large sample sizes and advances in computational approaches have allowed for phenome-wide studies of heritability on several million individuals from the United States.^8–10^ In these studies, genetic relationships have been inferred via family insurance plans or emergency contact forms. These approaches rely on strong assumptions and suffer from selection bias and confounding due to the selected nature of the populations. For example, vulnerable groups or patients with many comorbidities are less likely to obtain insurance, and the follow-up time in each study is rather limited. Another study, using the Danish pedigree, assessed the heritability for ten very broad disease categories and non-linear effects or shared environment were not taken into account.^11^ Hence, these estimates are likely to be inflated.

An extension of heritability is the genetic correlation that describes the degree to which two phenotypes share a genetic footprint.^3^ A significant correlation indicates that risk variants are in linkage disequilibrium, horizontal or vertical pleiotropy, and the correlation can be used to disentangle the genetic or environmental background of two correlated phenotypes.^12^ Large studies of genetic correlations have been used to identify previously unrealized similar disorders and suggest new nosologies.^8,13^ Furthermore, methodological advancements in statistical genetics have successfully utilized highly correlated traits to identify novel loci.^13,14^ Additionally, incorporation of genetically correlated traits in genomic prediction models may improve discrimination.^3,13^ Nonetheless, studies of genetic correlations are notoriously difficult because they require large sample sizes and necessitate comprehensive phenotype information with long follow-up to obtain accurate estimates.

The potential of assessing comorbid diseases temporally to extract the genetic component has been demonstrated in a limited number of autoimmune diseases.^15^ Furthermore, observational studies have shown that the order in which diseases manifest influences the comorbidity burden.^7^ The temporally ordered disease trajectories are therefore important to map to increase our understanding of disease progression and allow for patient stratification longitudinally.^6,15,16^ Thus, elucidating the contribution of genetics and environment to disease trajectories in a phenome-wide manner is important in a precision medicine setting to improve patient stratification.

Here, we perform a comprehensive phenome-wide study of the heritability and genetic correlations between diseases, disease pairs, and disease trajectories using nationwide data covering 6.3 million individuals over 40 years. This represents the most complete and detailed view of human disease heritability performed across an entire national healthcare scheme. We show that we can recapitulate previous studies but provide resolution for diagnoses at lower incidence, allowing more detailed exploration of the etiology of diseases at finer-grained calculations. We conduct an exhaustive analysis of sex-specific heritability. We also calculate comprehensive pairwise correlation of heritability of disease terms, illuminating the likely shared etiology between diseases, and explore the heritability of disease trajectories. In the paper, we highlight a number of observations from this comprehensive dataset, which are also available in summary form for every disease upon publication.

## Methods

### Ethical approval

The use of data from the MultiGeneration Registry-Lite was approved by the Danish National Archives (journal 19/04085). Informed consent or evaluation by a research ethics committee is not required for registry-based research in Denmark. The Danish Blood Donor Study (DBDS) was approved by the National Danish Committee on Health Research Ethics (1–10-72–95-13),the Zealand Regional Committee on Health Research Ethics (SJ-740), and the Data Protection Agency (P-2019–99). Genetic research within the DBDS was approved by the Danish National Committee on Health Research Ethics (1700407).

### Study population and setting

The nationwide observational population was identified using the Central Person Registry (CPR), MultiGeneration Registry-Lite (MGR-Lite), and LifeLines register at Statistics Denmark.^17^ Only the individuals present in all registries were included in the analysis. The population at risk was defined as those who were alive or born after January 1st, 1977, when the Danish National Patient Registry was established. The population at risk was followed until death, emigration, or the end of data (31^st^ December 2017) using the LifeLines register. Individuals who were not at risk were used to define relations in the data.

### Defining biological and non-biological relationships

We combined data from the CPR registry and MGR-Lite to create a unified Danish pedigree. The CPR registry records parent-child relations from children born 2^nd^ of April 1968 or hereafter. However, until 1978, relations were deleted when the child was 15 years of age. MGR-Lite is a registry that combines old versions of the CPR registry with information from digitized church books, thereby linking children born after 1953 and still alive in 1968 to their parents. If there were any disagreements between the two datasets, MGR-Lite was considered the trusted source. Furthermore, we removed relationships if the parental age was implausible (paternal age above 80 years, maternal age below 10 or above 64), and if the relationship was marked as an adoption.

### Validating biological relationships using genotype data

To assess the validity of the Danish pedigree we utilized genetic data from the Danish Blood Donor Study.^18^ In short, blood donors have been recruited since 2011. Currently, 92,861 individuals have been genotyped using the Infinium Global Screening Array from Illumina.^19^ Kinship was computed using a set of independent high-quality markers (excluding palindromic and non-autosomal markers, markers with MAF<1%, low call-rate (<99%) and markers in regions with high Linkage Disequilibrium) for the relatedness calculation, using KING.^20^ The Kinship coefficients estimated using genotyping data were compared to the theoretical kinship estimated from the pedigree, including only those relationships with a kinship coefficient > 0.085, corresponding to a relationship closer than first cousins.

### Outcomes

We performed a phenome-wide analysis of heritability across the ICD-10 codes, pairs, and trajectories. In the period 1977-1993 Denmark used ICD-8 codes, which were translated to ICD-10 using the mapping provided by Pedersen et al.^21^ We included all ICD-10 codes at the second, third, and fourth levels with a prevalence of at least 0.01%, and two related affected individuals. The latter was necessary because heritability could otherwise not be determined. Subsequently, we estimated the genetic correlations between pairs of diagnoses, where the heritability was at least 10%. Lastly, we also studied the heritability and genetic correlation of progression disease pairs, and the differences in genetic correlation between bidirectional disease pairs.^7^ We calculated disease pairs and disease trajectories using the approach defined in Jensen et al.^16^ using the full period 1977-2018. Progression disease pairs were defined as those with an increased risk and temporal directionality. Bidirectional disease pairs were defined as those that had an increased risk going both from A to B and B to A.^7^

### Statistical Analysis

Concordance between genotype and pedigree genetic relationships was calculated using the Spearman correlation. The 95% percentile confidence intervals (pCI) were estimated using bootstrapping (sampling with replacement, repeated 1,000 times). Estimating heritability and genetic correlations on the observed scale and then transforming it to the liability scale yields severely biased results when individuals are closely related (Supplementary Figure 1 and 2). Therefore, we estimated heritability and genetic correlations on the liability scale using a novel method scalable to full populations, yielding unbiased estimates of the heritability and genetic correlations on the liability scale (Supplementary Figure 3 and 4). In short, we minimized the difference between the expected product of the phenotypes versus the observed product, taking into account age and sex. This is similar to the Haseman-Elston regression cross-product method, but extended for binary phenotypes and implemented in Julia for computational efficiency. The credible intervals (CI) of the heritability and genetic correlation estimates were calculated by partitioning the population into ten subpopulations of roughly equal size and using the estimates to construct a credible interval. Members of the same family were ensured to be in the same subpopulation. As the largest extended pedigree included >5 million individuals, we partitioned the network into families of approximately 500 individuals. Covariance matrices describing additive genetic effects, non-additive genetic effects, and close environments were constructed using the same method and notation as described earlier^18^. For the non-additive genetic effects and close environments, we created covariance matrices for siblings, couples, and nuclear families, with all diagonal elements set to one. If two individuals were siblings, a couple, or in a nuclear family (siblings, couples, parent-children), the off-diagonal element was set to one. Owing to the extended pedigree design, it was necessary to limit each individual to only have one partner (to avoid negative definite covariance matrices), and include two nuclear family matrices, as a parent in one family can be a child in another. Posterior distributions of heritability and genetic correlations were estimated using a Bayesian beta regression model and are reported as 95% credible intervals (95% CI). A detailed description of this approach is available in Supplementary Methods. Heatmaps of genetic correlations were created using the R package ‘pheatmap’ (v1.0.12). Diagnoses were clustered using the complete hierarchical clustering method, with 1-correlation as the distance metric. Clustering disorder was calculated by iteratively identifying the nearest neighbor, and recording whether it was from the same ICD10 chapter. Confidence intervals were determined using a two-sided binomial test, and the 95% confidence interval (95% pCI) was reported.

## Results

### Data overview

We assembled the extended Danish pedigree, consisting of 6.3 million individuals, starting from the full Danish population of >8.8 million individuals utilizing nationwide registries (Figure 1A). Using a novel methodology, scalable to large populations, heritability and genetic correlations were estimated from the extended pedigrees across the full ICD-10 ontology (Figure 1B). Prior to applying our pedigree-breaking algorithm, the average size of the extended pedigrees was 18. After breaking the pedigree, the largest family had 762 members. The average follow-up time was 28.7 years. For individuals born prior to 1977, 60% (2,366,134) had 40 years of follow-up during the period 1977-2017. For individuals born after 1977 we had complete follow-up from 1977-2017 for 79.2% (1,830,262) (Supplementary Figure 5). Hence, the cohort is a general representation of the Danish population, with long and near-complete phenotypic information. We validated the pedigree relationships using a sample of 92,861 Danish blood donors and found that pedigree relationships were concordant with the calculated genotype relationships (Spearman *ρ*= 0.788, 0.783; 0.794 95% pCI).

**Figure 1.**
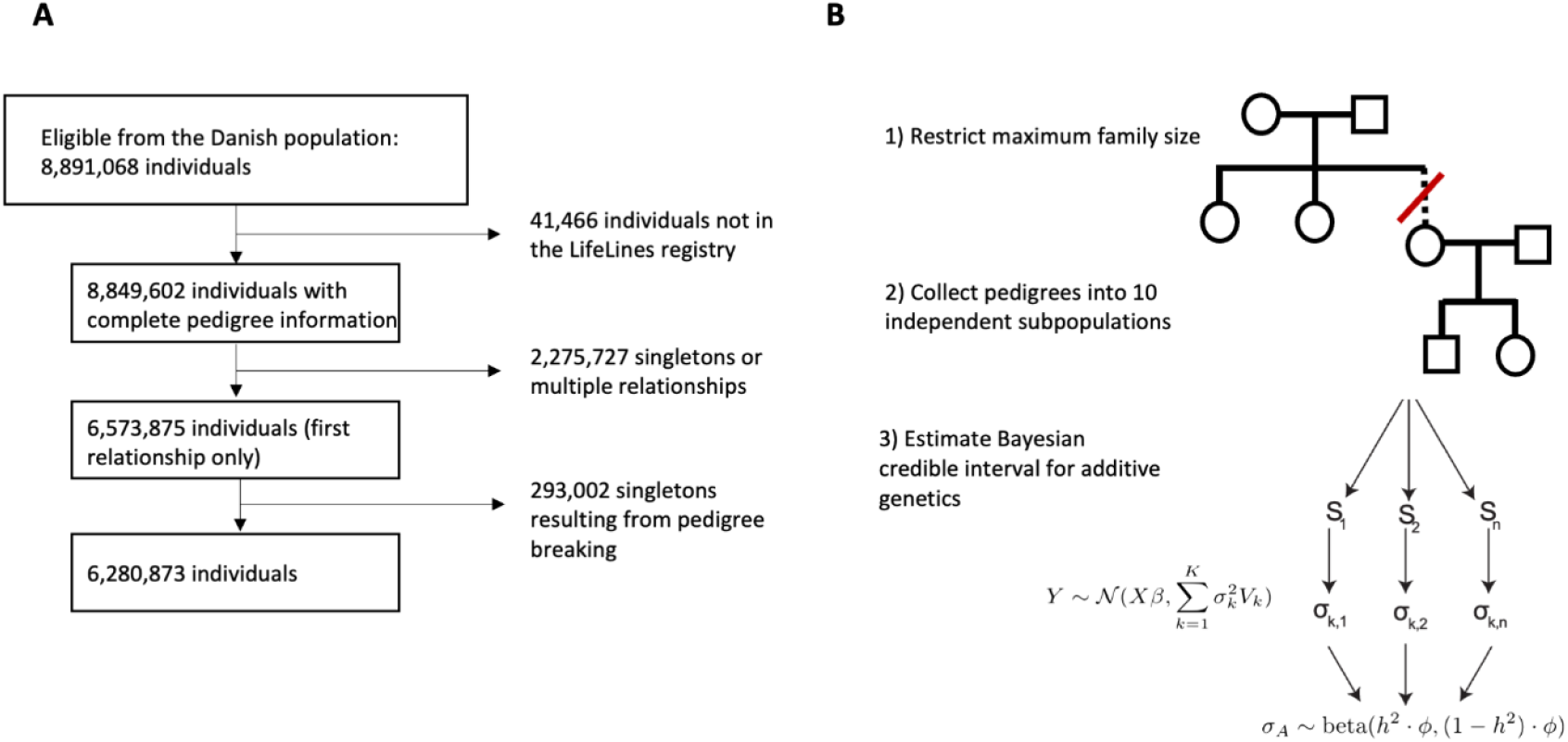
(A) Flowchart of how individuals were selected, starting from the full Danish population of >8.8 million individuals. (B) Workflow for determining heritability and genetic correlations. 1) The pedigree is broken into smaller families of ∼500 individuals. 2) For each of the 10 independent subpopulations, the heritability or genetic correlation is determined. 3) The credible interval is estimated using a Bayesian model.

### Heritability and genetic correlation across the full spectrum of diseases

We estimated the additive genetic heritability (h^2^) across 866 level three ICD-10 codes fulfilling the selection criteria, adjusted for non-linear genetic effects and close environment. Across the disease areas, congenital malformations, endocrine diseases, and mental disorders had the highest heritability (Figure 2). The ICD-10 codes with the highest heritability were diseases with known large genetic components, often with well described specific genes known where genetic variants are known to impact diseases, such as cystic kidney disease (Q61, h^2^=0.85, 0.78; 0.89 95% CI), osteochondrodysplasias (Q78, h^2^=0.77, 0.66; 0.83 95% CI), and congenital malformation syndromes (Q87, h^2^=0.75, 0.62; 0.82 95% CI). Looking beyond these known genetic diseases, almost 50% (N = 424 codes) of the level three ICD-10 codes displayed a heritability with a magnitude of h^2^ > 0.1. Resolving the heritability at the less specific 2nd level (N=183 codes) and more specific 4th level (N=1,519 codes) of ICD-10 supported the same pattern of heritability (Supplementary Figure 6 and 7) with 63% (N=971) Level 4 codes having h^2^ > 0.1, indicating the importance of acquiring accurate family history for diagnoses, even outside of well-established genetic conditions. Heritability estimates across the full ICD-10 ontology are available in the Supplementary Materials.

**Figure 2:**
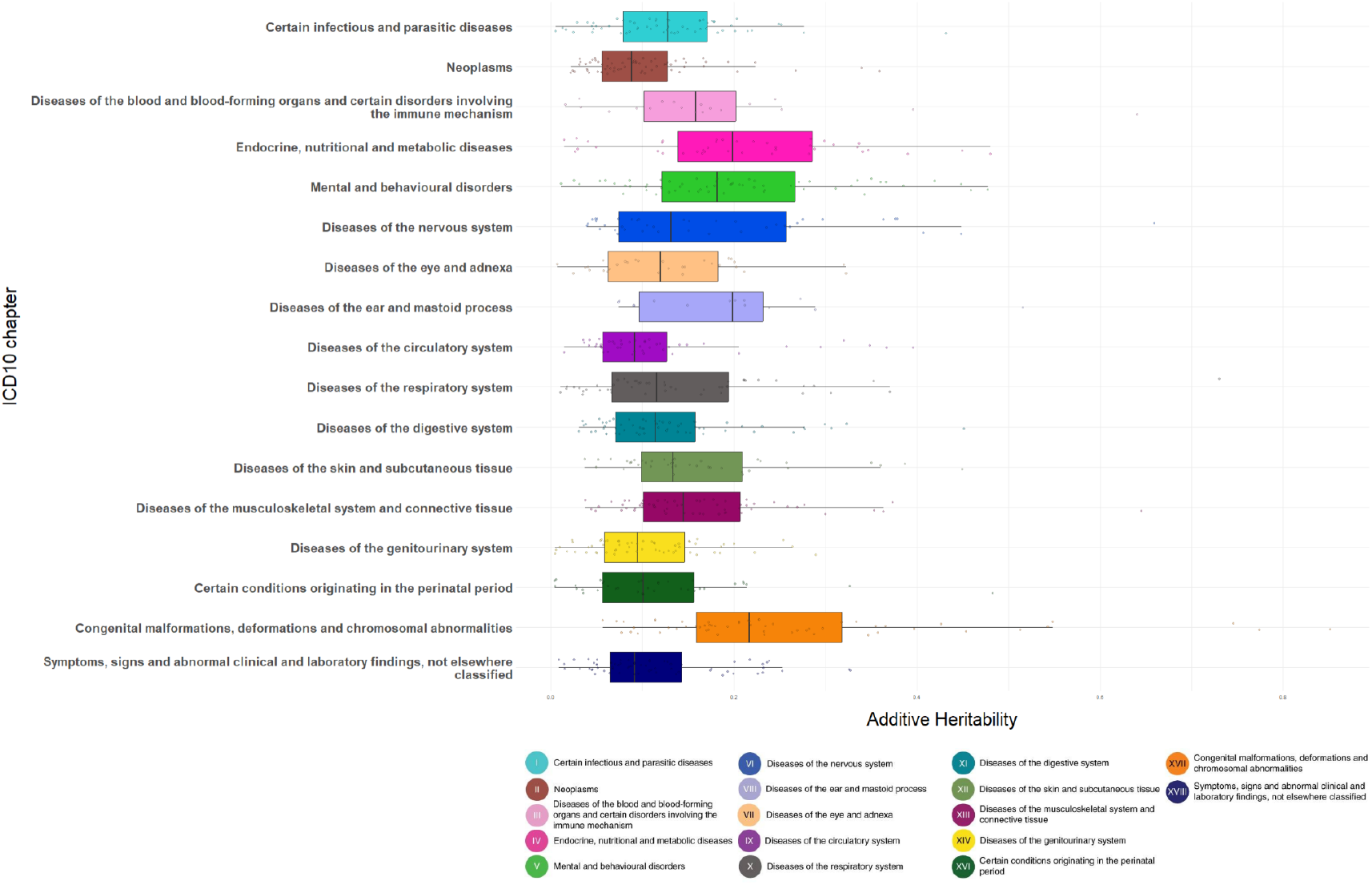
Estimated additive heritability across 866 ICD10 codes at the third level. The most heritable disease areas were congenital malformations, endocrine diseases, and mental disorders. ICD10 codes are grouped and colored according to the chapter they belong to.

### Comparison with prior studies

We compared our findings with three previous studies assessing the heritability of many phenotypes using pedigree or twin data: two from the United States of America (pedigree-based) and one from the Scandinavian countries (twin-based). Earlier studies only included diseases with a higher prevalence, owing to small sample sizes. Nonetheless, estimates of heritability were consistent, with correlation coefficients ranging from 0.33-0.57 or 0.16-0.67, depending on whether environmental variance components were included (Supplementary Methods).

### The confounding of shared environment and non-additive genetic effects

Shared environment is an important confounder of additive heritability estimates. Comparing a model including environmental components (sibling, spousal, and nuclear family relationships) to a model that includes only additive genetics provides clear evidence, with a higher heritability estimate when disregarding the environment (Figure 3). Diseases pertaining to infection, perinatal conditions, and the endocrine system showed the highest level of bias. “Acute upper respiratory infections” and “Viral infection of unspecified site” were the only infections not significantly affected by the close environment, suggesting a common underlying disease genetic susceptibility. Investigating the spousal component (Sp), we found that OCD (Sp=0.57, 0.51; 0.62 95% CI), HIV infection (Sp=0.52, 0.46; 0.58 95% CI), and Gonococcal infection (Sp=0.35, 0.29; 0.42 95% CI) had the highest proportions of explained variance. This is in line with current knowledge, namely that assortative mating is present amongst individuals with OCD.^22^ Consequently, disregarding environmental components can severely inflate the heritability estimates for many diseases. However, for diseases such as cardiovascular and pulmonary disorders, the estimates appear robust and unaffected by environmental confounding in this study.

**Figure 3:**
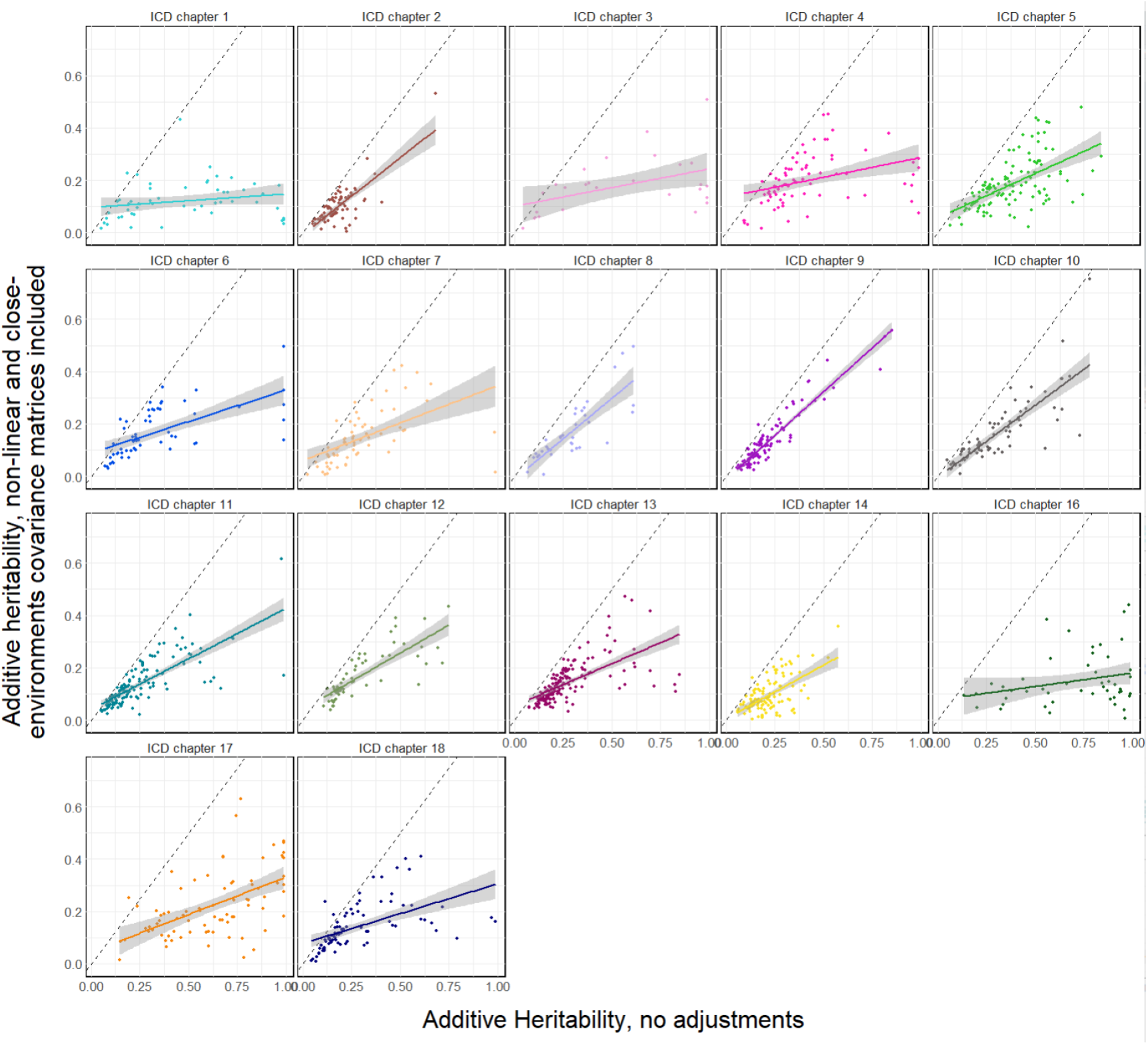
Comparing the additive heritability in a model with only the additive genetics variance component (x-axis) versus a model including the additive genetics, sibling, spousal, and nuclear family variance components (y-axis). In gray the 95% confidence interval for the linear model. The heritability is biased upward when not accounting for the environment. Color coding follows the same scheme as in Figure 2.

By extending the model to include a dominance effect (D), we observed an inverse relationship between the magnitude of additive and dominance effects (Figure 4). There was no contribution from dominance when an additive effect was present and vice versa. For instance, myeloid leukemia had no notable additive heritability (h^2^=0.06, 95% 0.03; 0.14). However, there was a large attributable fraction of variance to dominance heritability (D=0.42, 95% 0.36; 0.48). This most likely reflects that certain familial cases, such as mutations in the *CEBPA* gene, follow a dominant inheritance pattern.^23^ The dominance component primarily captures the variance from the residual and sibling components (Supplementary Figure 8).

**Figure 4:**
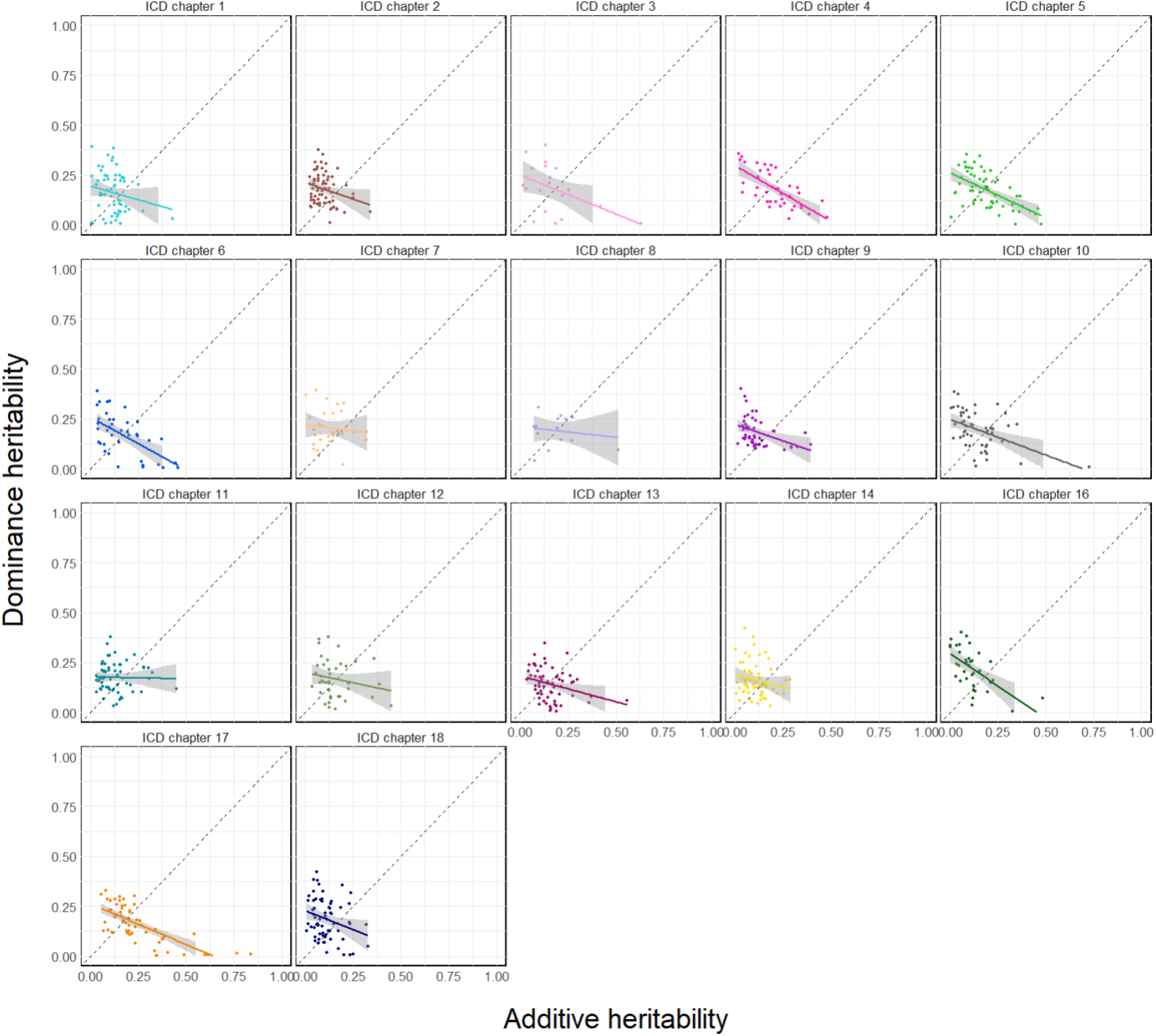
Comparing the additive heritability (x-axis) and dominance (y-axis) in a model that includes sibling, spousal, and nuclear family variance components. There is an inverse relationship between the effect from additive heritability and dominance. Color coding follows the same scheme as in Figure 2.

### Resolving phenotypic heterogeneity across ICD-10

The phenotypic heterogeneity of the different ICD-10 levels has not been systematically described. We compared estimates of heritability between the second, third, and fourth level of ICD-10; these different levels represent a mixture of phenotypic subtypes of diseases in some cases, and in other cases, useful groupings of disorders that affect similar physiological systems or other rationales for collections. As expected, we observed that estimates of heritability, in general, increased with diagnosis specificity (i.e., higher heritability at higher levels of ICD-10). For example, heritability estimates for the level 2 block “Other soft tissue disorders” were relatively low (h^2^=0.19, 0.19; 0.20 95% CI). However, with increased specificity, heritability estimates increased; at the third level of codes related to fibroblastic disorders, heritability increased (h^2^=0.28, 0.24; 0.32 95% CI), and at the fourth level, heritability for Dupuytren was even higher (h^2^=0.47, 0.44; 0.50 95% CI). Similarly, the heritability of fibromyalgia was also notably higher (h^2^=0.46, 0.37; 0.55 95% CI) (Figure 5A). There were a handful of cases where the opposite pattern was also seen, for example, in Cleft lip and Cleft palate diseases (Figure 5B). Here the level two ICD-10 codes for Cleft lip and Cleft palate have a high heritability (h^2^ = 0.49, 0.36;0.61 95% CI), whereas the more precise child terms in levels three and four show lower heritability, e.g., Cleft lip, unilateral (h^2^ = 0.30, 0.25; 0.37 95% CI). Additional examples regarding viral hepatitis, myoneural disorders, and systemic atrophies are provided in Supplementary Figure 9.

**Figure 5:**
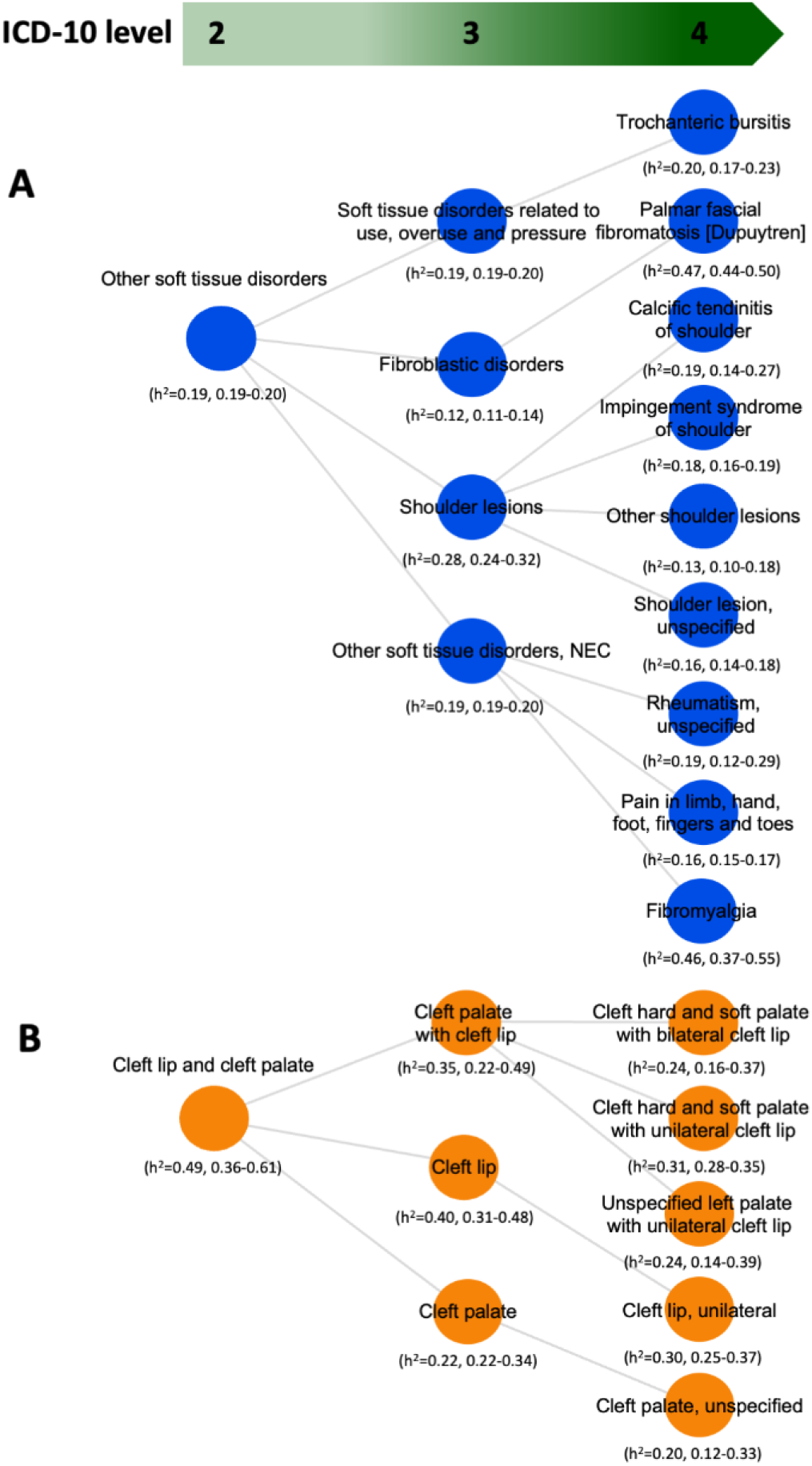
Heritability at different levels of ICD10 within the “Haemolytic anaemias” and “Cleft lip and cleft palate” blocks. Haemolytic anaemias and Cleft lip and cleft palate decrease in heritability when resolved into more specific phenotypes.

### Sex-specific estimates of heritability and genetic correlations

We compared heritability between men and women across 500 ICD-10 codes, which had a prevalence of at least 0.1% in both, and found 74 cases in which the heritability differed (Figure 6). Some of the largest differences were observed in hypertrophy of breast, bone disorders, and scoliosis. Generally, heritability was higher in women (69/74 disorders), which aligns with previous genetic studies.^24^ Deviations in heritability could be attributed to sex-linked biological differences, differences in disease prevalence, or differences in the environment between men and women (e.g., hormones, diet, or work environment).

**Figure 6:**
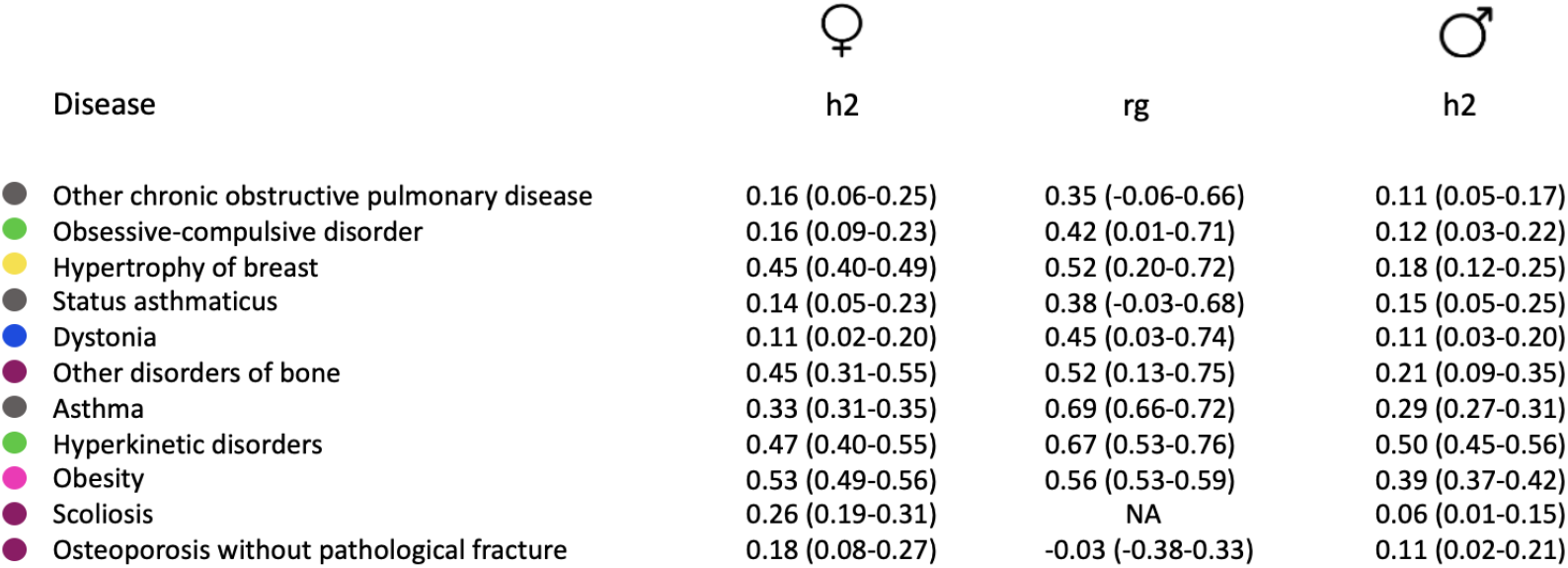
Comparing heritability and genetic correlations between men and women for selected phenotypes. Several disorders, such as breast hypertrophy, OCD, and obesity displayed remarkable sex differences.

To evaluate whether the underlying etiology is shared between men and women, we considered the genetic correlation (rg) between the same disease in men and women. Here, a genetic correlation of less than one implies non-overlapping pathways in disease initiation or interactions with the environment not captured. For 37 of the 500 investigated diagnoses the genetic correlation was < 0.8. This included diagnoses with a previously described sexual dimorphism, such as obesity (rg=0.56, 0.52; 0.59 95% CI), hypertrophy of breast (rg=0.52, 0.19; 0.72 95% CI), and asthma (rg=0.69, 0.66; 0.72 95% CI). However, pronounced sexual dimorphism was also observed in obsessive compulsive disorder (rg=0.42, 0.01; 0.71 95% CI), hyperkinetic disorders (rg=0.67, 0.53; 0.76 95% CI), and dystonia (rg=0.44, 0.03; 0.74 95% CI). For 15 diagnoses, we could not rule out a genetic correlation of zero. This included diseases where we have previously described a difference between men and women, such as osteoporosis without fracture (rg=-0.025, -0.37; 0.33 95% CI), status asthmaticus (rg=0.38, -0.03; 0.38 95% CI), and chronic obstructive pulmonary disease (rg=0.35, -0.05; 0.65 95% CI).^6^ Out of the 500 diagnoses investigated, there were none with an inverse correlation between men and women.

### Genetic correlations

We investigated the pairwise genetic correlations (rg) of 424 level three ICD-10 codes with a heritability of > 10%. This resulted in 89,031 estimates of genetic correlation, of which only 3% (2,635) had an inverse correlation, 42% (37,537) had a positive correlation, and 55% (48,859) had no correlation. 645 failed to reach convergence using our methodology (see Methods). The diagnoses did not cluster systematically by the assigned ICD-10 chapter (Figure 7). This indicates that while ICD-10 is grouped based on anatomical and functional groups, they do not necessarily have a common genetic basis. There were two large clusters that contained a number of mental disorders. One included bipolar disorders, schizophrenia, substance abuse, and hepatitis, suggesting vertical pleiotropy (Fig 8A). The other contained mental disorders in childhood and adolescence, epilepsy, and microcephaly (Fig 8B). Another highly correlated cluster consisted mostly of infectious diseases, localized in distinct anatomical locations, such as influenza, hordeolum, and impetigo, but from different ICD-10 chapters (Figure 8C). A cluster related to migraine, headaches, and unspecified pain across the body was also identified (Figure 8D). Examining the discordance, that is, the proportion of closest neighbors not from the same chapter, we found that the diagnoses from the chapters “Diseases of the blood and blood-forming organs” (0%, 0%; 29% 95% jCI), “Diseases of the eye and adnexa” (0.6%, 0.0%; 0.32% 95% pCI), and “cancer” (22%, 0.6%; 48% 95% pCI) were least likely to cluster with a diagnosis from the same chapter. Mental disorders nearly always formed intra-chapter clusters (88%, 74%; 96% 95% pCI). All other chapters had very similar estimates. Interestingly, at the fourth ICD10 level, “Cancers” displayed a higher degree of order (57%, 35%; 77% 95% pCI) (Figure 9). Looking further than just the first neighbor, “Cancers” quickly saturated towards a high degree of order. “Diseases of the blood and blood-forming organs” remained highly disordered (Supplementary Figure 10).

**Figure 7:**
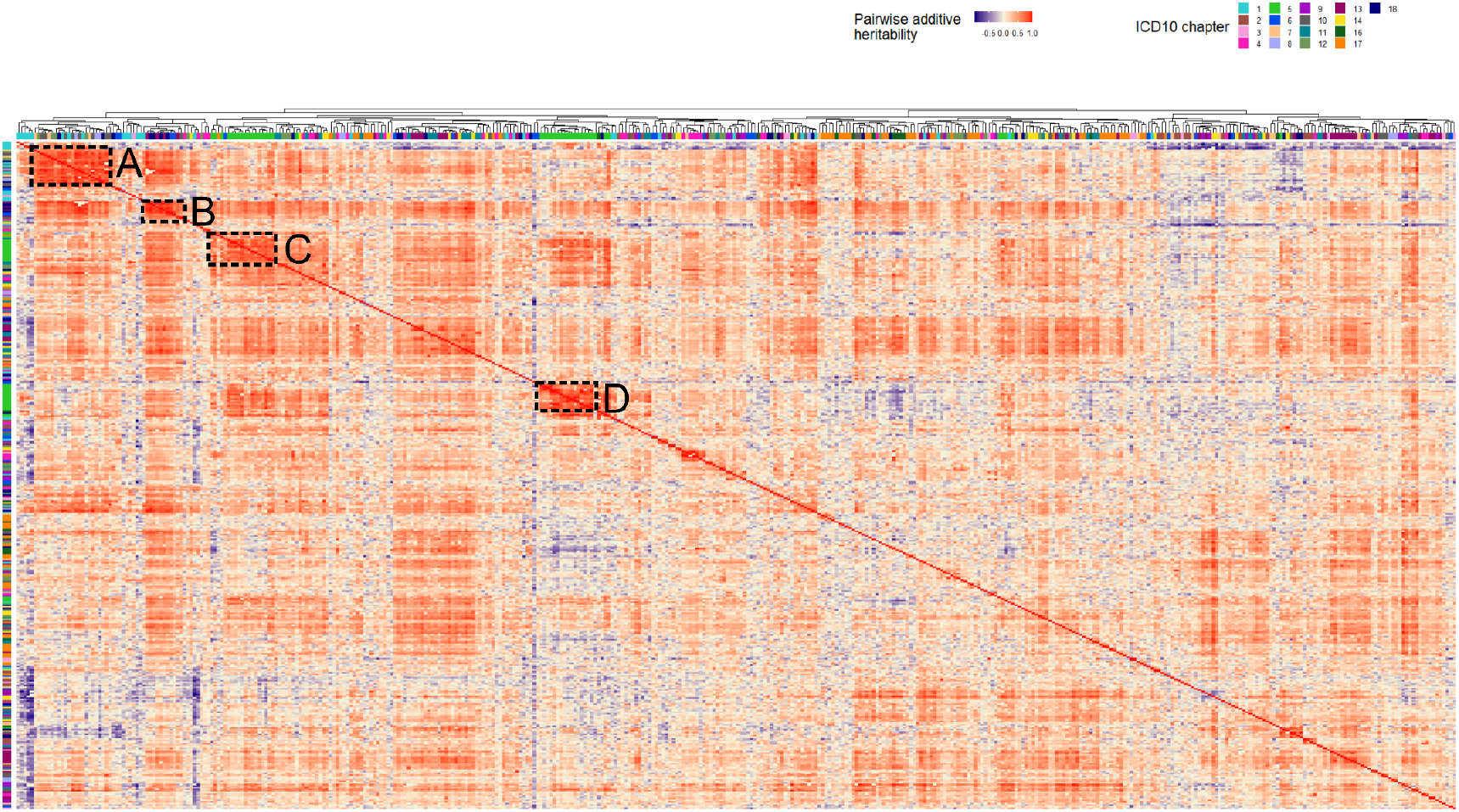
Clustering of ICD10 level three codes based on estimated genetic correlation. Letters refer to sub-plots in Figure 8. Coloring of the dendrogram refers to the ICD10 chapters.

**Figure 8:**
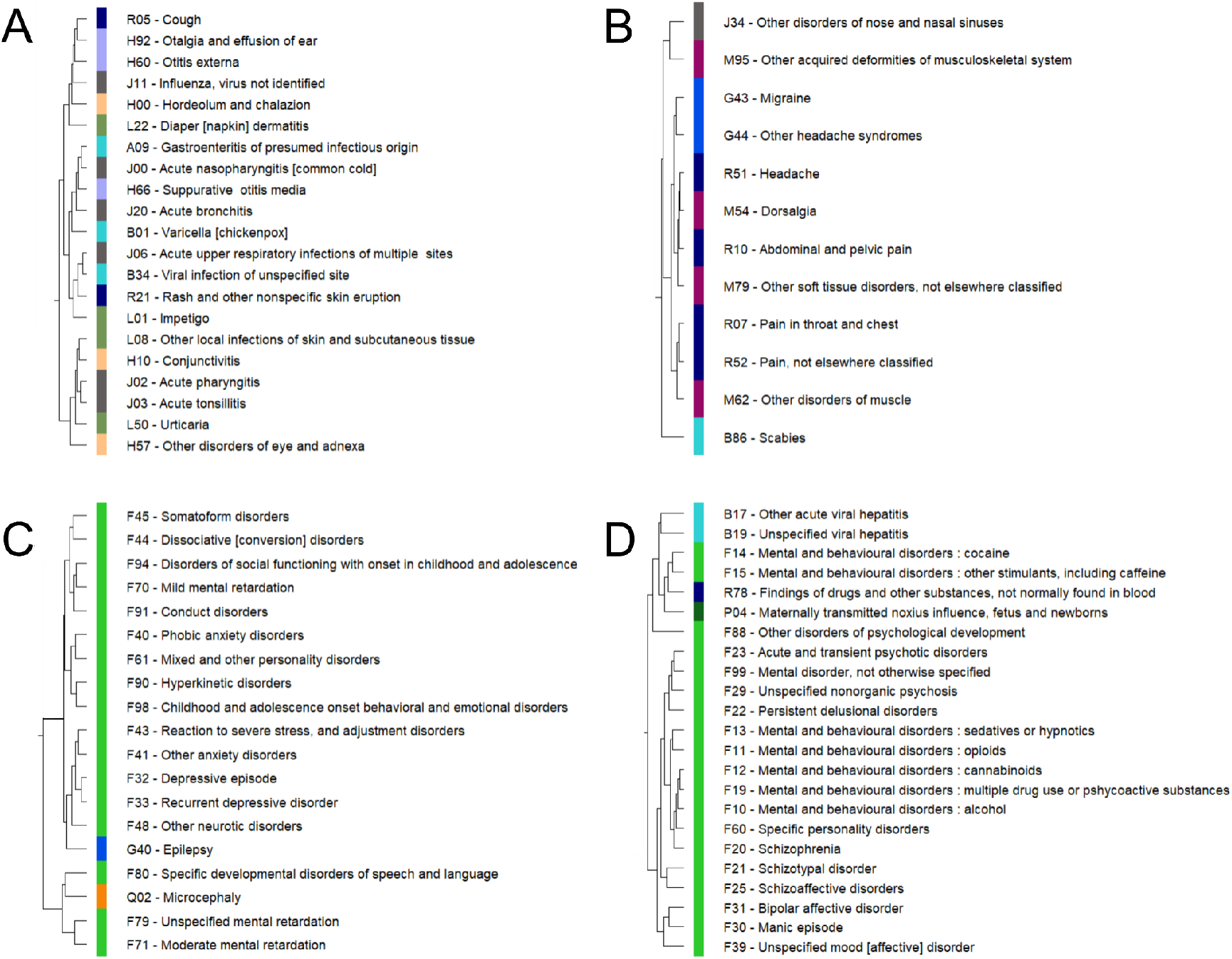
Selected clusters from the heatmap. Numbers refer to the selected clusters in Figure 7.

**Figure 9:**
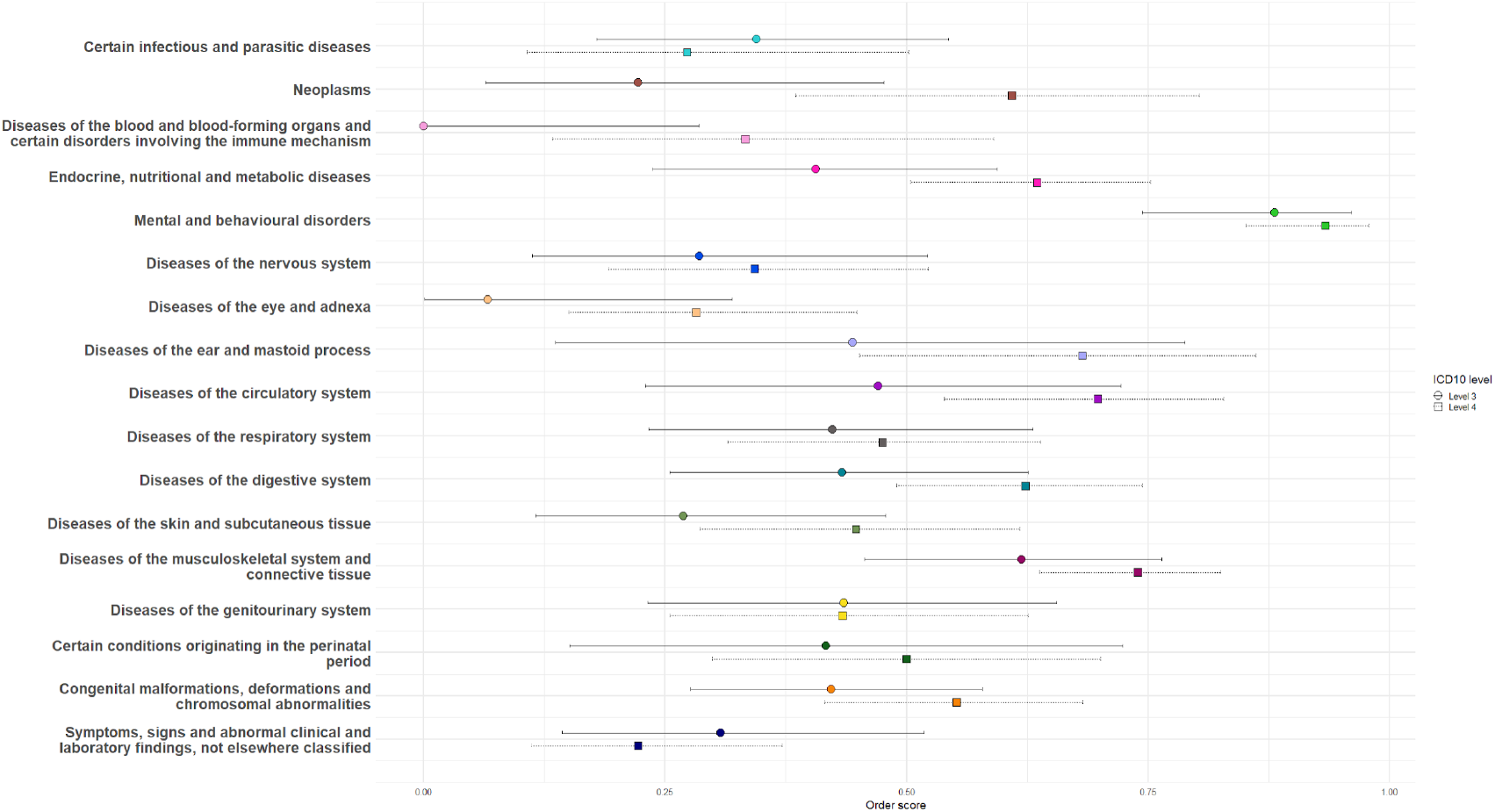
Disorder across ICD10 chapters, based on the nearest neighbor from the dendrogram in Figure 7.

Focusing on cardiology, dilated and hypertrophic cardiomyopathy (DCM and HCM, respectively) appeared genetically distinct (rg=-0.09, -0.40; 0.24 95% CI), fitting the known genetic etiology of these diseases. Nonetheless, we found very similar estimates of genetic correlations with other diseases such as atrial fibrillation, cardiac arrhythmias, and palpitations (Figure 10).

**Figure 10:**
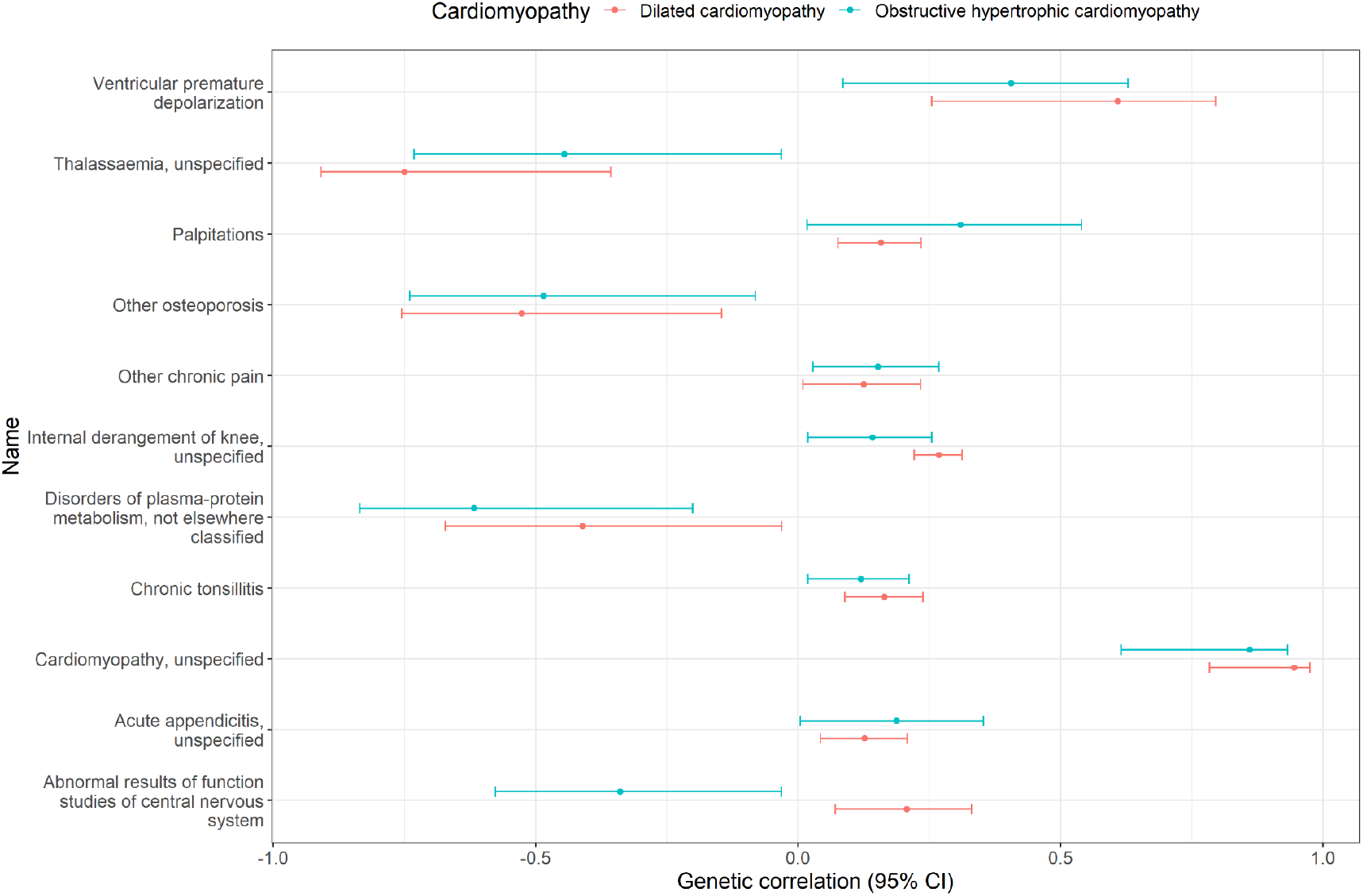
Comparing diseases genetically correlated to both dilated and hypertrophic cardiomyopathy.

With regard to diabetes, we observed that hyperplasia of prostate and Parkinson’s disease were both inversely correlated with diabetes, which observational evidence has hinted towards due to the protective effects of diabetes medication.^25,26^ The etiology of fibromyalgia is not understood, and here we show genetic correlations with multiple diseases (N = 311 ICD-10 level four codes). Overall, genetic correlations were positive. Nonetheless, malignant cancers (C43.5, C44.4, C44.5) of the skin showed an inverse correlation. Clotting related disorders (“Hereditary deficiency of other clotting factors” and “Thrombocytopenia”) also displayed an inverse correlation. Further studies on these diseases could shed light on the etiology of fibromyalgia.

### Heritability of bidirectional associations

We identified 6,518 cases in which there was a bidirectional relationship between diagnoses, that is, disease A increased the risk of disease B, and vice versa. Of these, 320 pairs had a heritability of > 10%, resulting in 160 estimates of genetic correlation. Largely, the majority of bidirectional relationships had a common genetic background (117/160 bidirectional pairs) (rg > 0.85). This indicates that often bidirectional relationships represent the same disease, implying that there is an underlying biological process, such as vertical pleiotropy, and that the order of expression is arbitrary and possibly determined by the environment or other non-additive genetic factors.

Bidirectional relationships worth noting were: Recurrent depressive disorder and Other functional intestinal disorders (rg=0.54, 0.18-0.77 95% CI); and, Asthma and GERD (rg=0.55, 0.19; 0.78 95% CI) (Figure 11).

**Figure 11:**
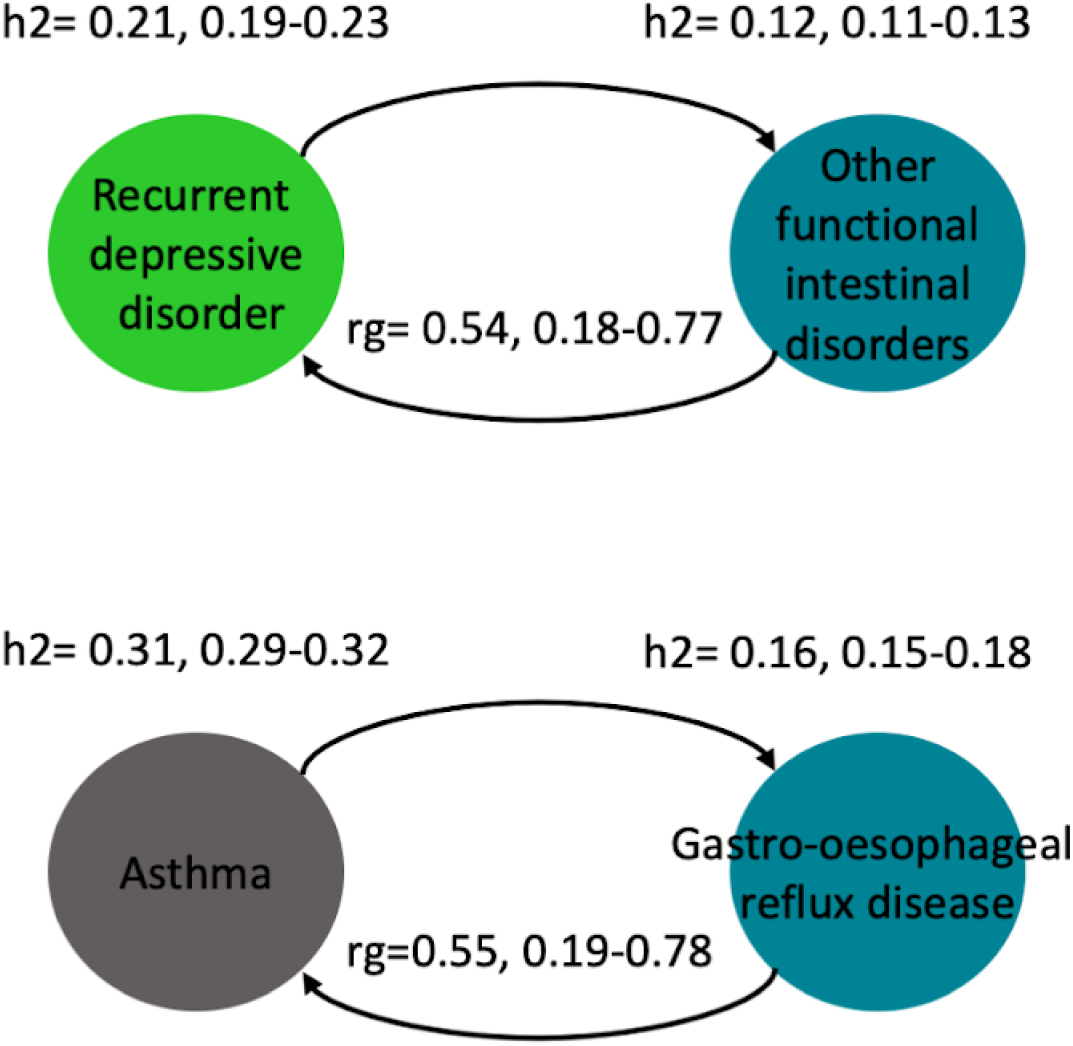
Bidirectional heritability. Two examples of disease pairs where the order of manifestation had a different underlying genetic liability.

### Heritability of disease pairs and disease trajectories

We identified 1,216 disease pairs with a significantly increased relative risk and directionality occurring in at least 0.1% of the population (Methods). Of these, 380 disease pairs had two or more affected family members. Eight trajectories of length 3 had a prevalence greater than 0.1%, and at least two affected family members. We found three trajectories with a heritability of > 0.2 (Figure 12B). Many trajectories seem to represent a clinical process in deducting a classification arising during clinical investigations or complex diagnostic pathways leading to broad categories (e.g., “general pain”). An interesting exception is trajectories that start with a diagnosis of Type II diabetes, but switch to Type I diabetes, consistent with a relatively constant incidence of Type I diabetes over time, which is easier to misclassify in middle age to Type II diabetes.^27,28^ There are two such trajectories discovered in this work leading to retinal issues respectively. This emphasizes the importance of considering a Type I etiology of diabetes at all ages of glucose metabolism presentation. This may be a result of clinical investigations, for example, not taking family history into account, but this could also reflect inaccuracies in coding practice.^29^

**Figure 12:**
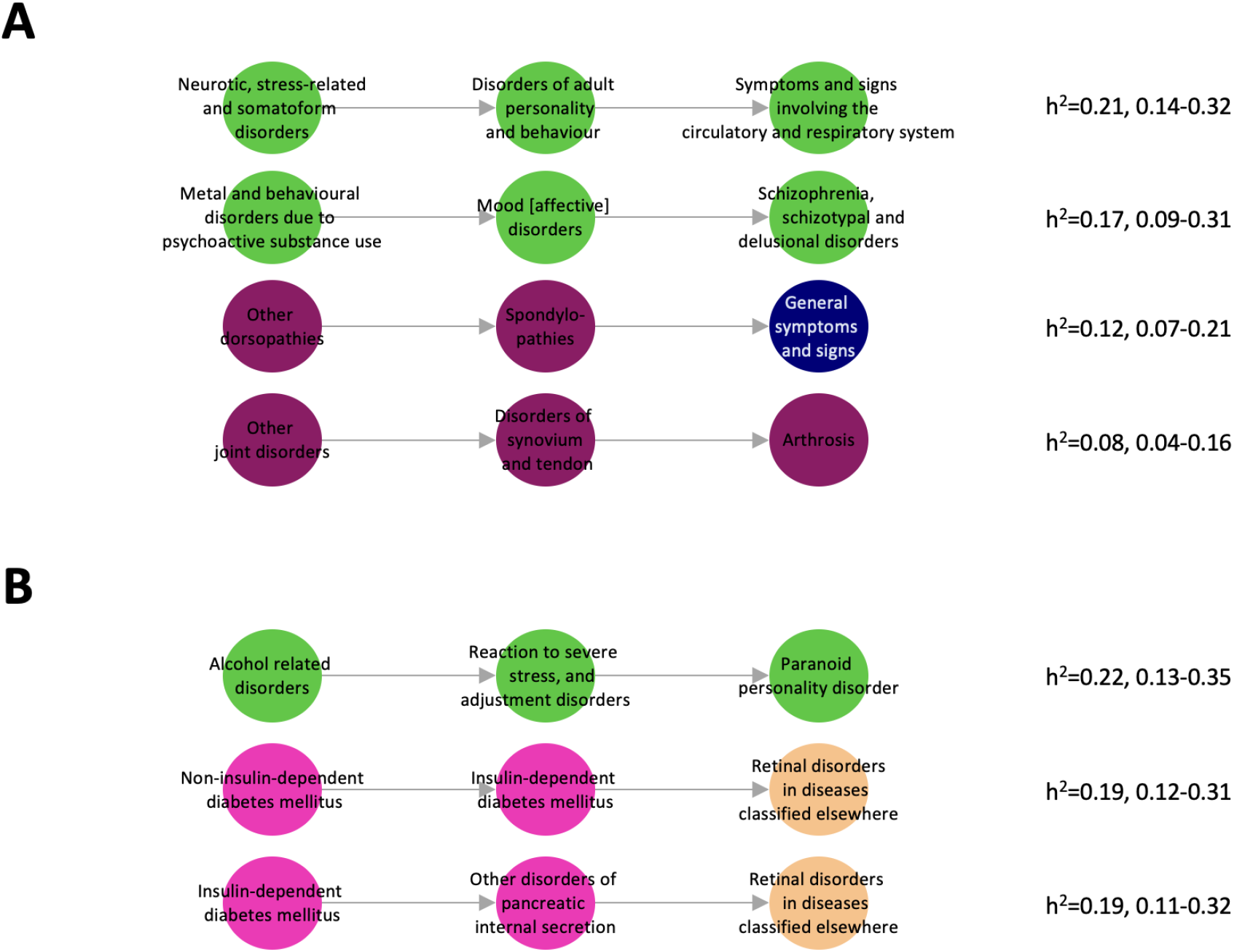
Heritability of disease trajectories. (A) Trajectories at the 2nd ICD10 level. (B) Top 3 trajectories at the 3rd ICD10 level. Nodes are coloured according to chapter, following the same color scheme as in Figure 2.

At the 2nd ICD-10 level, the trajectory with the highest heritability described the transition from “Neurotic, stress-related and somatoform disorders”, to “Disorders of adult personality and behavior”, and finally “Symptoms and signs involving the circulatory and respiratory systems” (h^2^=0.21, 95% CI 0.14; 0.32) (Figure 12A). Furthermore, we also identified a trajectory starting from “Mental and behavioral disorders due to psychoactive substance use”, to “Mood [affective] disorders”, and finally “Schizophrenia, schizotypal and delusional disorders” (h^2^=0.17, 95% CI 0.09; 0.31).

## Discussion

### Summary

We present a comprehensive phenome-wide study with in-depth characterization of the genetic determinants of disease, disease progression, and disease trajectories. The underlying data foundation was based on a large nationwide cohort. Despite the relatively short generational depth, dense interconnectivity can be established through blood or marriage. Coupled with detailed complete hospital admissions and a long follow-up period, we were able to systematically examine diseases, comorbidities, and disease trajectories across the ICD-10 ontology. Here, we provide evidence of the underlying genetic components of disease comorbidities and trajectories.

### Limitations and Strengths

This study used a nationwide cohort with follow-up from as early as 1977, which included all age groups in a homogenous cohort. There was no inherent bias in representability owing to the nationwide setting. Nonetheless, the analysis of diseases relied on secondary use of hospital data, and we acknowledge that some conditions are underrepresented or poorly coded as they are diagnosed and/or treated outside secondary care. Along the same line, there could be systematic errors present we are not aware of that could induce sporadic correlations between diseases^30^. Additionally, heritability may change over time, and it is important to note that these phenotypes have been collected over the last 40 years when disease prevalence, health seeking behavior and diagnostic practices have changed. Furthermore, we use a single term to represent the environmental correlation between siblings, spouses, and nuclear families. It is a strong assumption that this captures both the environmental and non-additive genetic effects of dominance and assortative mating. Although not perfect, not taking this into account yields heritability estimates biased upward. Additionally, assortative mating results in inflated genetic correlations^31^. Here, we consider this through the spousal variance component and show that our findings are in line with the literature. Future studies should explore new ways to integrate information from extended families into environmental covariance matrices. Errors in data on family relationships could bias results, but we show using a cohort of blood donors using genotyped data, this problem is likely to be very minor. The population-wide design and long follow-up allowed us to study both rare and late-onset diseases. Nonetheless, studying the heritability of disease progression and trajectories is difficult, as it requires an old population with known ancestors due to the time it takes to accumulate diseases to form a trajectory.

## Conclusion

The broad theme of this work is consistent with previous systematic studies and clinical research, such as the existence of some strongly heritable diseases, the presence of sex-biased diseases with heritable components, and the shared, common etiology of many diseases. In our exploration of this dataset, we found a number of novel relationships between specific genetic diseases and subphenotypes such as cleft-palate disease, the trajectories of diabetes diagnoses and the correlation patterns of mental disease, but we are confident that there are many more discoveries present in this dataset to be cross-referenced or contrasted with known clinical knowledge. The results presented here should be taken into consideration in all disease studies when designing and interpreting genetic and epidemiological results. We provide comprehensive results for the entire community to be readily used. The results are widely applicable to the design and utility of genome-wide association studies, shared etiology of diseases, and genomic risk prediction, and add to our understanding of the fundamentals of disease development.

## Data Availability

All data produced will be made available online at https://h2.cpr.ku.dk

## Acknowledgements

The study was supported by funding from the Novo Nordisk Foundation (grant agreements NNF14CC0001 and NNF17OC0027594). The work was also carried out as a part of the BRIDGE Translational Excellence Programme (bridge.ku.dk) at the Faculty of Health and Medical Sciences, University of Copenhagen, funded by the Novo Nordisk Foundation (NNF18SA0034956).

