## Supplementary for "Uncovering the Heritable Components of Multimorbidities and Disease Trajectories: A Nationwide Cohort Study"

### Population wide heritability of disease courses: a nationwide cohort study

Westergaard et al

#### Supplementary Methods

##### Estimating heritability and genetic correlations on the liability scale

Traditionally, the observed heritability ($h_o^2$), estimated using e.g. REML or Haseman-Elston regression, is transformed to the liability scale ($h_l^2$) using a result derived from normal distribution theory,

$$h_l^2 = \frac{P*(1-P)}{\phi(t)^2} * h_o^2$$

In which $P$ is the population prevalence, and $t$ is the threshold. However, this linear relationship breaks down in the presence of closely related individuals[1], and results in a severe bias of results as the heritability increases (Supplementary Figure 1). Similarly, estimates of genetic correlations are also affected as the absolute magnitude increases (Supplementary Figure 2).

An alternative is to employ Bayesian models with a probit link that naturally models the binary nature of the data, such as those implemented in MCMCglmm or THRGIBBS. These models do not scale well to large populations (we estimated a runtime of 14 days/phenotype). Instead, we developed a novel and scalable approach to estimate heritability on the liability scale. We assume a liability threshold model. For a population of size N, in which each individual is indexed $i$ ($i \in \{1, ..., N\}$) and has the phenotype $Y_i$,

$$Y_i = \begin{cases} 1 & \text{if } \ell_i > 0 \\ 0 & \text{if } \ell_i <= 0 \end{cases}$$

where $l_i$ is the liability, and distributed as,

$$\ell \sim \mathcal{N}(X\beta, \sum_{k=1}^{K} \sigma_k V_k)$$

in which $K$ denotes one of the $k$ covariance components in the model, $\sigma_k$ is the k'th variance component, and $V_k$ is the k'th covariance matrix. Furthermore, it is assumed that $\sum_{k=1}^{K} \sigma_k = 1$ to ensure identifiability. X is the design matrix, $X \in \{0, 1\}^{NxJ}$, where we assume each row sums to one, i.e. each person belongs to one group. The $j$ groups are strata of year of birth (in decades), stratified by sex. If there are J groups, there will be J fixed effects, denoted $\beta_j$. Let $p_j$ denote the prevalence in group $j$. Because we assume $\sum_{k=1}^{K} \sigma_k = 1$, we can estimate the fixed effect for person $i$ belonging to group $j$,

$$p_i = P(Y_i = 1) = P((X\beta)_i + N >= 0) = P(N <= \beta_j) =>$$

$$\phi(\beta_j) => \beta_j = \phi^{-1}(p_j)$$

Under the further assumption that $N \sim N(0, 1)$. We use the reparametrization $\sigma_k^{\ 2} = \frac{\theta_k}{\sum_k \theta_k}$ for $\theta \in [0, 1]$. This ensures that $\sum_{k=1}^{K} \sigma_k = 1$. Let $\theta := (\theta_1, ..., \theta_K)$. Then, we minimize the loss function,

$$L(\theta) = \sum_{j<i} (E_\theta(Y_i Y_j) - Y_i Y_j)^2$$

Because we estimated the fixed effects beforehand, $E_\theta(Y_i\ Y_j)$ can be calculated efficiently as a double integral (see Appendix). Also, because only have J different fixed effects and many entries in the matrices reappear many times, we need to calculate significantly less than $\frac{N(N-1)}{2}$ different expected values to evaluate the loss function.

The approach can also be extended to estimate the genetic covariance, which can be used to calculate the genetic correlation, ρ, between two outcomes, A and B, as follows,

$$\rho(A, B) = \frac{cov(A,B)}{\sqrt{h_A^2 * h_B^2}}$$

Simulation results using one of the subpopulations (N ~ 620.000) indicate that the method can recover additive heritability values reliable down to a prevalence as low as 0.001%, when including sibling and spousal covariance matrices (Supplementary Figure 3). The empirical bias is 0.0036 and the mean absolute error is 0.034. Simulations of genetic correlations indicate that the model provides unbiased results (Supplementary Figure 4).

#### Covariance matrices for additive genetics, non-additive genetics, and close environment

The additive- and dominance- genetic relatedness matrix was created using the R package nadiv (2.17.2), from the extended pedigree of 6.3 million individuals. Following the approach from Gary-McGuire et al. and Bochud et al. we define four covariance matrices to model the non-additive genetics and close environment, namely a matrix describing siblings, couples, and nuclear families. We restricted each individual in the pedigree to only have one partner. Otherwise, modeling the couple and nuclear family relationships would become unnecessary complex. Nonetheless, as we are working with extended pedigrees, two covariance matrices were needed for the nuclear family component, since an individual can be a part of two nuclear families (see figure below).

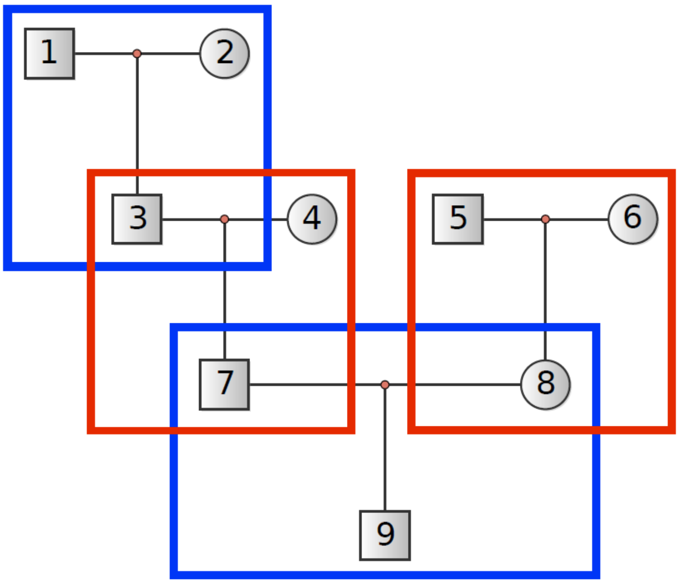


If two individuals were siblings, a couple, or in a nuclear family (siblings, couples, parent-children), the off-diagonal element was set to one (see example below)

$$S = C_{\text{Sibling environment}} = \begin{array}{c} \text{Father} \\ \text{Mother} \\ \text{Child}_1 \\ \text{Child}_2 \end{array} \begin{pmatrix} 1 & 0 & 0 & 0 \\ 0 & 1 & 0 & 0 \\ 0 & 0 & 1 & 1 \\ 0 & 0 & 1 & 1 \end{pmatrix}$$

$$C = C_{\text{Couple environment}} = \begin{array}{c} \text{Father} \\ \text{Mother} \\ \text{Child}_1 \\ \text{Child}_2 \end{array} \begin{pmatrix} 1 & 1 & 0 & 0 \\ 1 & 1 & 0 & 0 \\ 0 & 0 & 1 & 0 \\ 0 & 0 & 0 & 1 \end{pmatrix}$$

$$F = C_{\text{Family environment}} = \begin{array}{c} \text{Father} \\ \text{Mother} \\ \text{Child}_1 \\ \text{Child}_2 \end{array} \begin{pmatrix} 1 & 1 & 1 & 1 \\ 1 & 1 & 1 & 1 \\ 1 & 1 & 1 & 1 \\ 1 & 1 & 1 & 1 \end{pmatrix}$$

$$I = C_{\text{Individual environment}} = \begin{array}{c} \text{Father} \\ \text{Mother} \\ \text{Child}_1 \\ \text{Child}_2 \end{array} \begin{pmatrix} 1 & 0 & 0 & 0 \\ 0 & 1 & 0 & 0 \\ 0 & 0 & 1 & 0 \\ 0 & 0 & 0 & 1 \end{pmatrix}$$

The modeling of the environment in pedigree data is highly underdeveloped. Nonetheless, as we show, not taking the environment into account leads to an inflation of the heritability estimates.

#### Credible intervals for heritability and genetic correlations

Credible intervals for estimates of heritability were estimated using a Bayesian model, where we assume that the value estimated from each of the ten subsamples, denoted $\sigma_A$ follow a beta distribution, which is naturally bounded in the interval (0, 1) and well suited to model the proportion of variance explained. The beta distribution is parametrized in terms of the mean, $h^2$, and precision, $\phi$.

$$\sigma_A \sim beta(h^2 \cdot \phi,\ \left(1 - h^2\right) \cdot \phi)$$

In which $h^2$ and $\phi$ are modeled on the logit and exponential scale, respectively,

$$h^2 = logit(h_0^2)$$

$$\phi = exp(\phi_0)$$

To complete the model, we assign priors on the coefficients,

$$h_0^2 \sim t(7,\ -1,\ 1)$$

$$\phi \sim N(0,\ 1)$$

Since there are no covariates in the model, the prior on $\phi$ has virtually no impact on the estimates of $h^2$. The prior on $h^2$ concentrates a lot of the probability mass close to zero, thus being a conservative prior due to the a priori expectation that there is no heritability and regularizes estimates with large variance.

Estimates of genetic correlations, denoted ρ, were transformed to be in the interval (0, 1), as follows

$$X = \frac{p+1}{2}$$

And the same beta regression model was applied, with the only exception of a prior concentrating mass around 0.5, corresponding to a genetic correlation of zero,

$$h_0^2 \sim t(7,\ 0,\ 1)$$

Estimates from the Bayesian models were then transformed back to the original scale, [-1, 1],

$$p = (X \cdot 2) - 1$$

All intervals are reported as the 95% Bayesian Credible Intervals (CI). Models were fit using rstanarm[3], running for 10,000 iterations across 4 independent chains. The first half, 5,000, were used for tuning parameters of the Hamiltonian Monte Carlo No-U-Turn sampler. Convergence was assessed by calculation of R-hat values and number of divergences. Only if all R-hat values were < 1.01 and there were no divergences had the model converged.

#### Supplementary Results

##### Comparison with prior studies

We compared our findings with three previous studies assessing the heritability of many phenotypes using pedigree or twin data: two from the United States of America (pedigree based), and one from the Scandinavian countries (twin based). Estimates of heritability and genetic correlations were consistent, with correlation coefficients ranging from 0.33-0.57 or 0.16-0.67, depending on whether environmental variance components were included or not (Figure 1, Table 1). We note, however, that the earlier studies only included diseases with a higher prevalence, due to small sample sizes.

The study of Wang et al.[4] relied on nuclear families, using similarly structured variance components to describe the environment as in this paper, but differences in methodology makes direct comparison of results difficult. For instance Wang et al. pools the environmental variance, but this results in the conflation of genetics and environmental effects due to genetic dominance and assortative mating. The Wang et al. study limited the population to those who had been in the population the longest, which may itself be a bias, as that requires a continuous healthcare insurance with the same company over time.

The study by Polubriaginof et al.[5] used electronic medical records. However, they neglected accounting for non-additive effects by only including a maternal effect. As we show, the sibling and spouse environment heavily confounds the heritability and skews it upwards. Indeed, this is also evident from the halving of the correlation coefficient between the models (Table 1). Furthermore, their proposed method, which in theory can scale to large populations, cannot be used to estimate genetic correlations.

Athanasiadis et al.[6] estimate heritabilities using Falconer's Method, which does not take into account the close environment that is shared by near relatives. As we show, this leads to substantial upward biased estimates of heritability. Furthermore, diseases are grouped at the chapter level of ICD-10, which are internally very heterogeneous in terms of prevalence and genetic/environmental causes.

#### Supplementary Figures

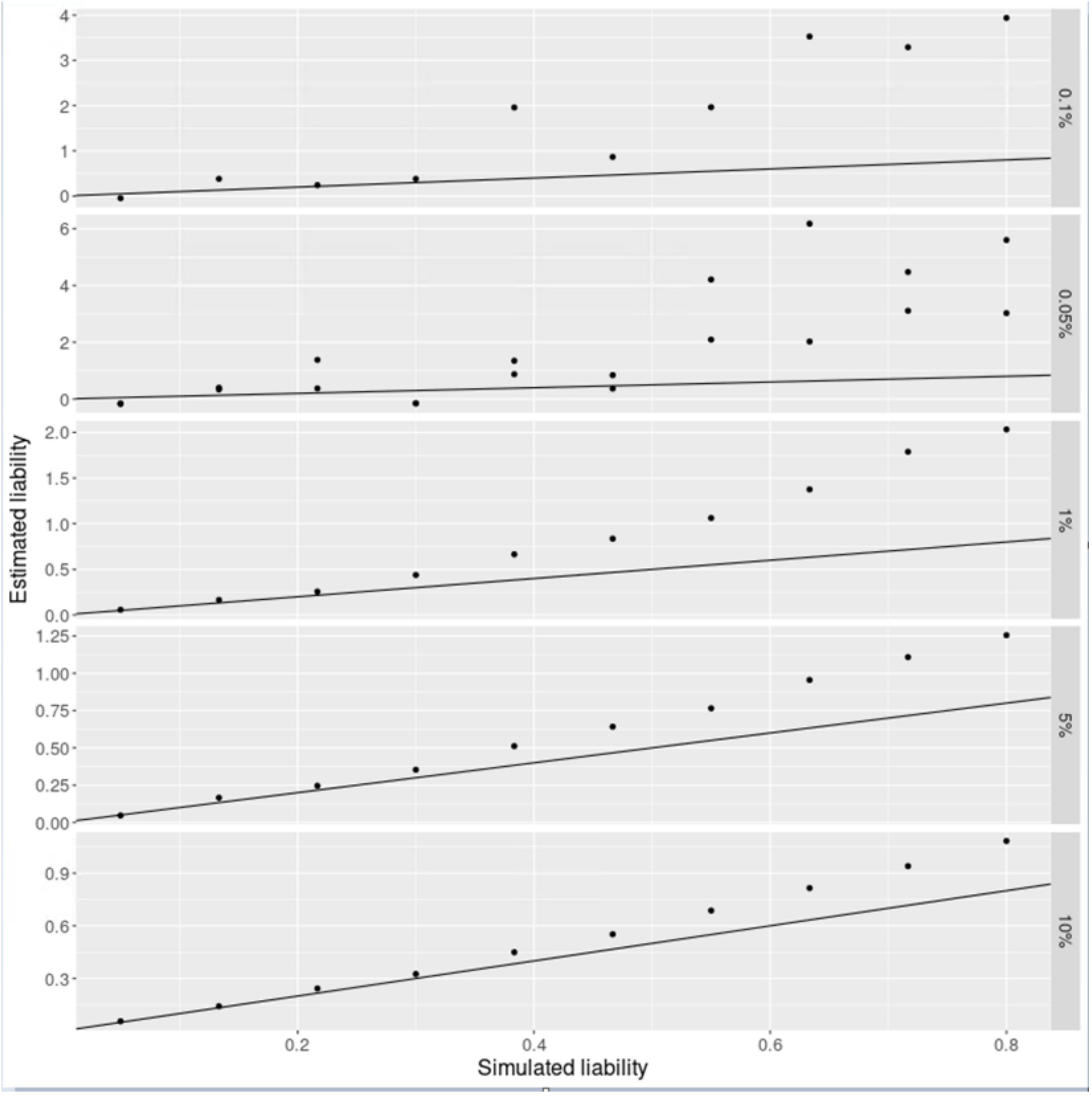


Supplementary Figure 1: Simulated heritability on the liability scale versus estimated liabilities using Eq. X.

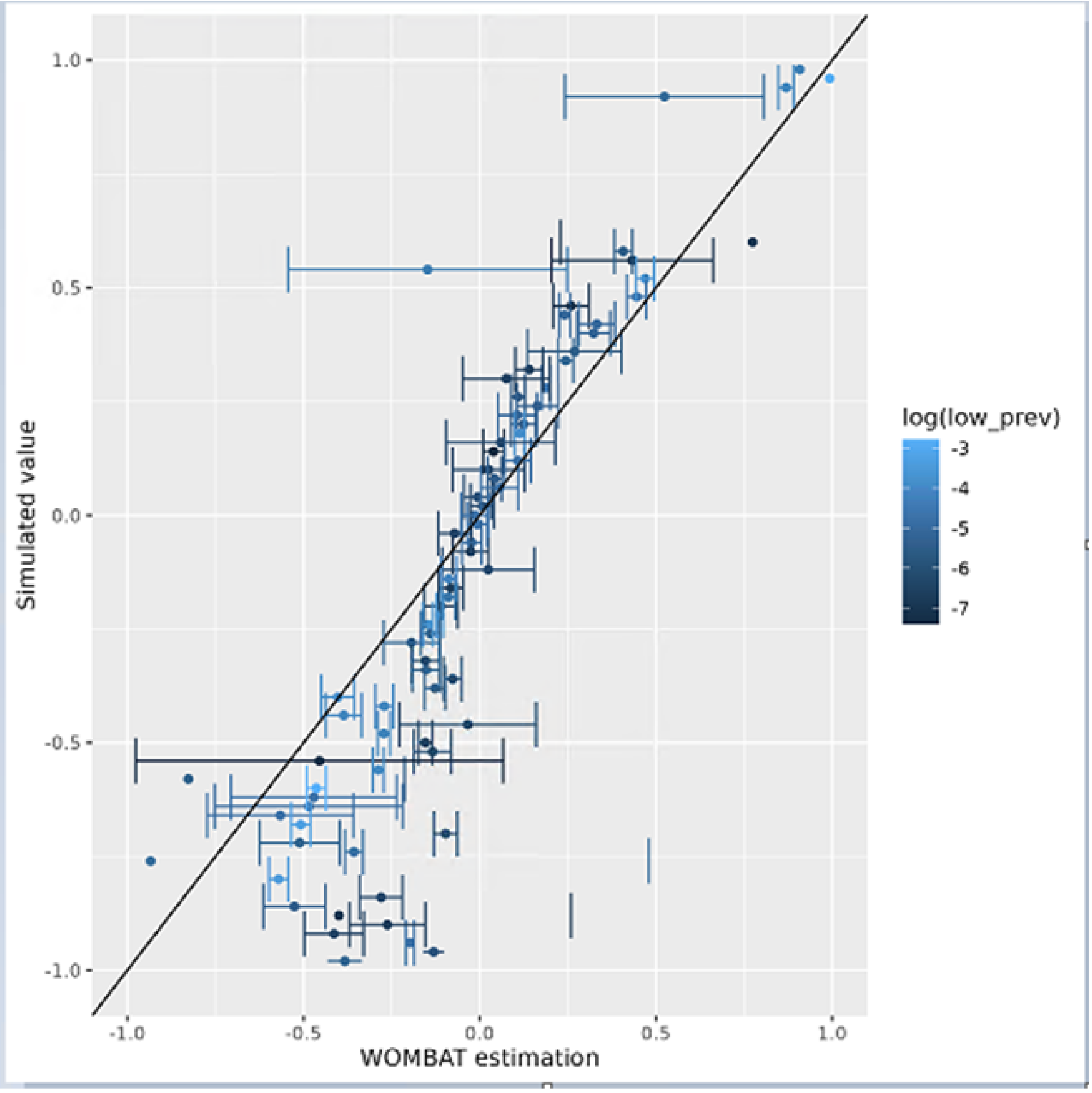


Supplementary Figure 2: Estimates of genetic correlations simulated from binary phenotypes using the mixed model equation approach implemented in WOMBAT.

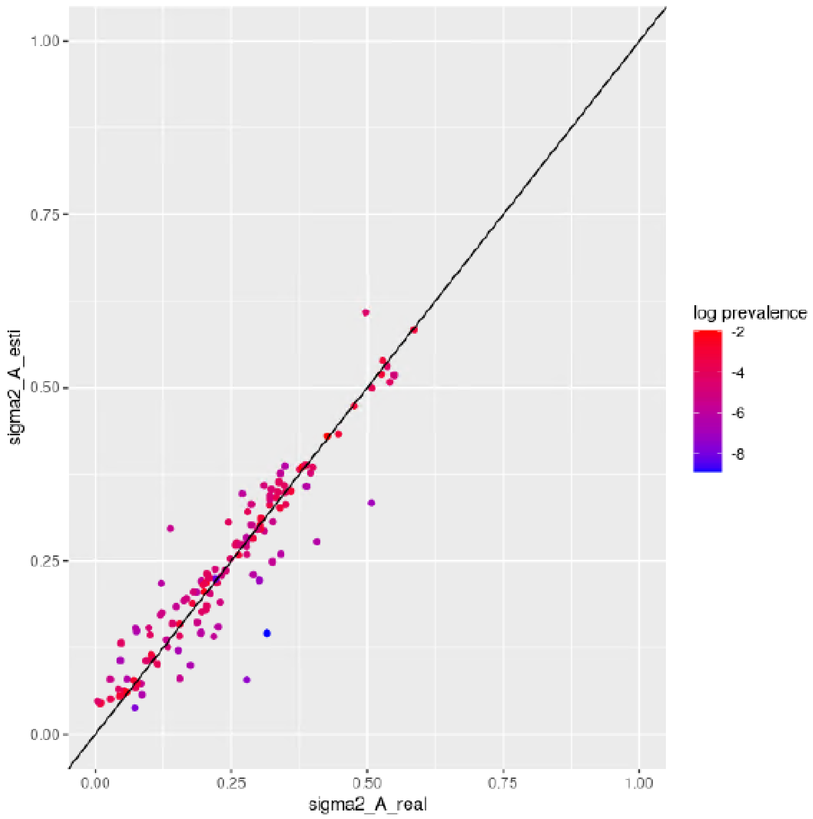


Supplementary Figure 3: Simulation of 129 phenotypes with prevalences ranging from 0.0003 to 0.13. Values are simulated uniformly across the 1-norm unit circle.

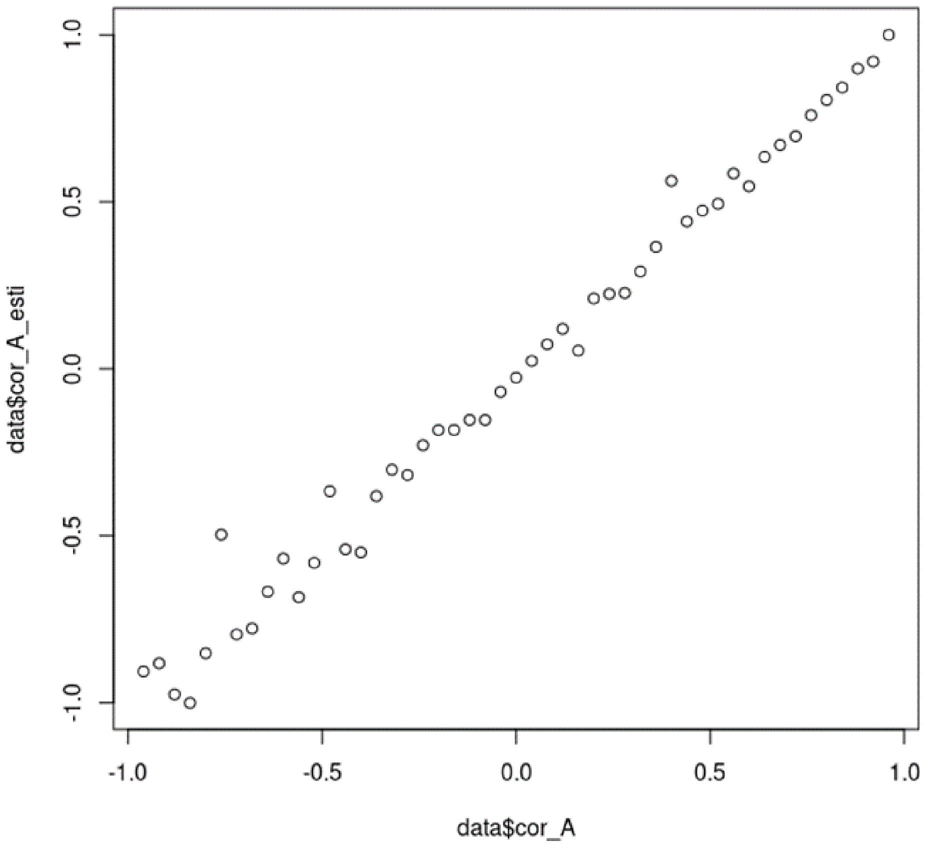


Supplementary Figure 4: Simulated values of genetic correlations

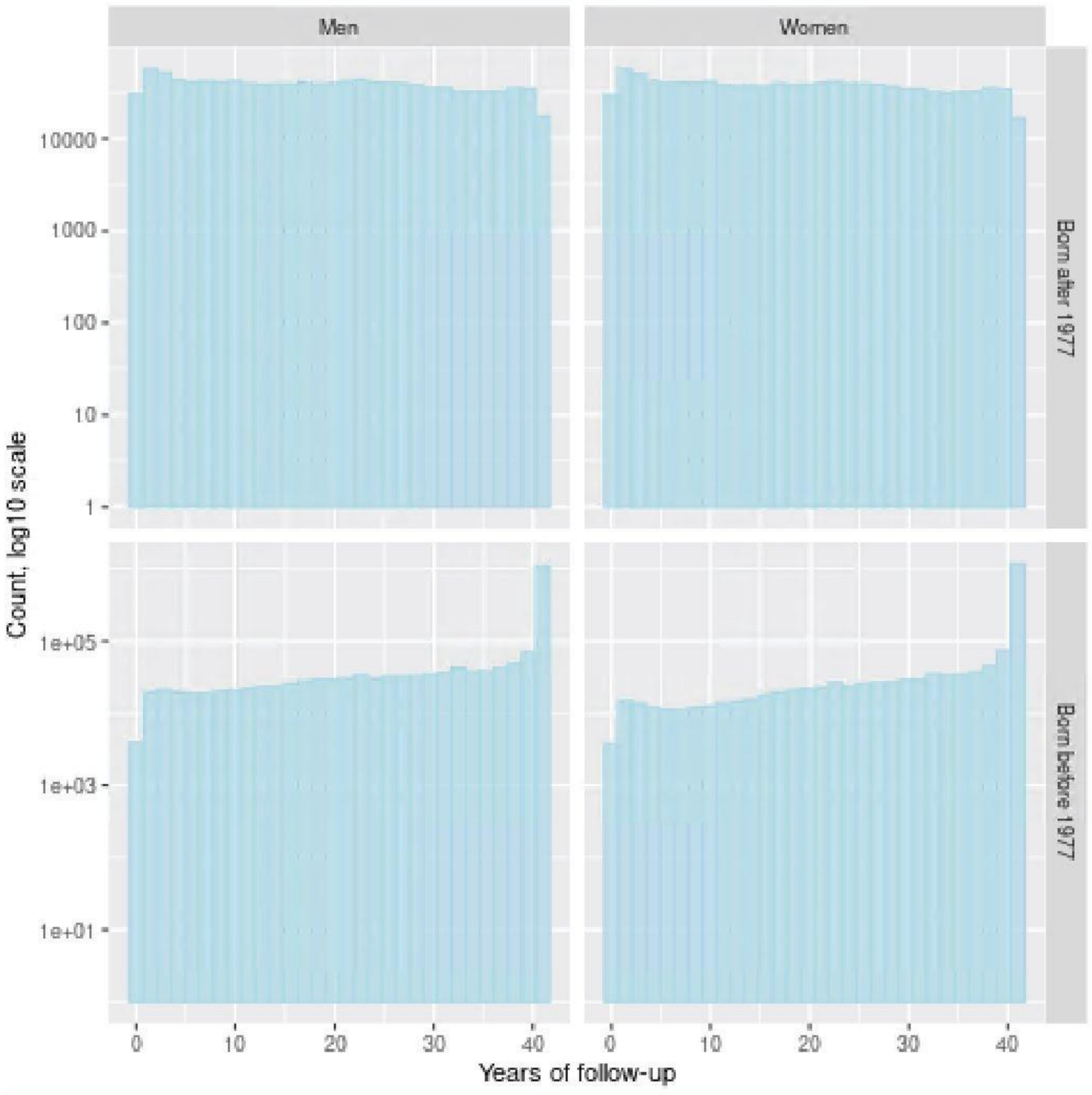


Supplementary Figure 5: Distribution of follow-up time in the Danish population, stratified by sex and year of birth. Phenotypic data from the Danish National Patient Registry was available from 1977 and onwards.

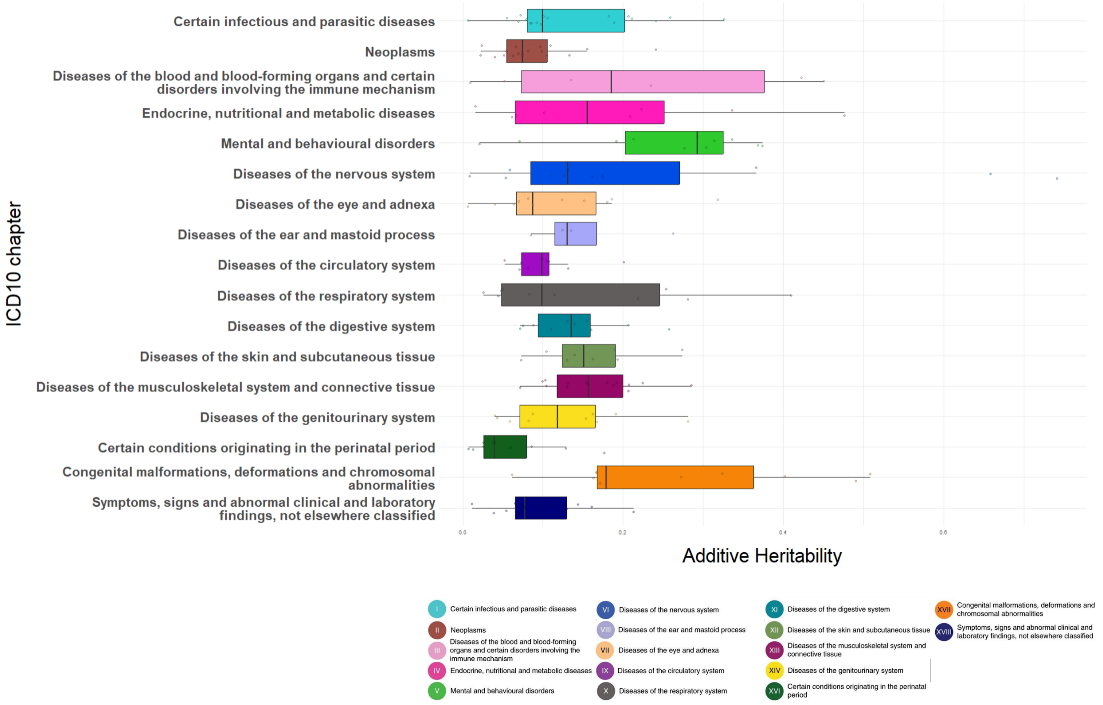


Supplementary Figure 6: Heritability of 2nd level ICD-10 codes.

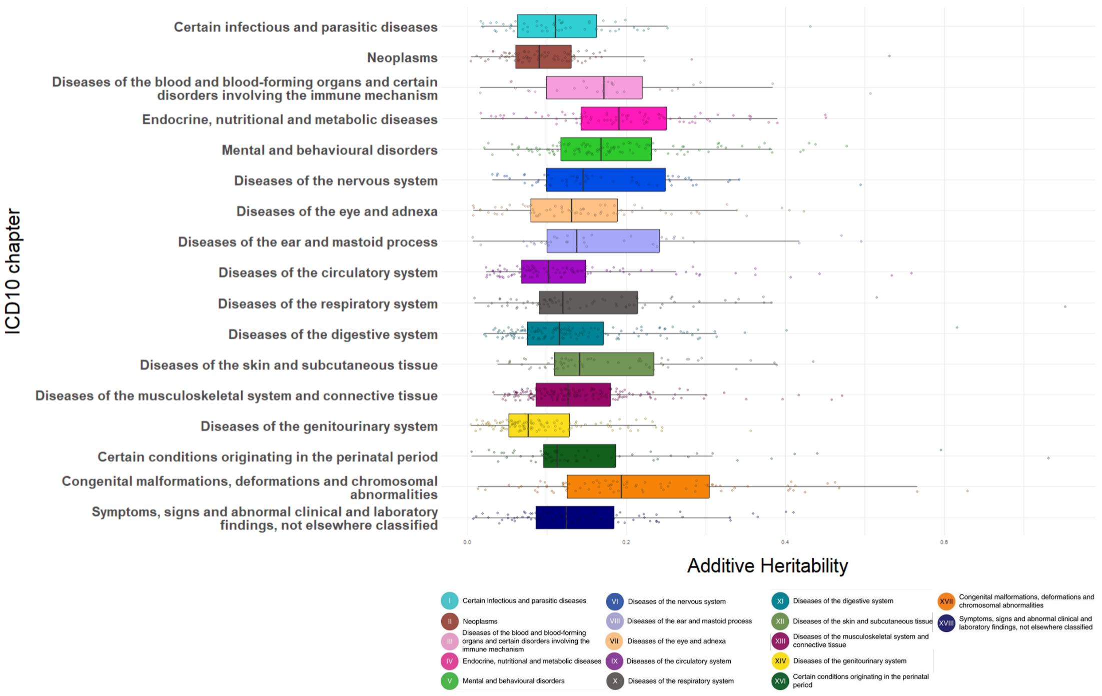


Supplementary Figure 7: Heritability of 4th level ICD-10 codes.

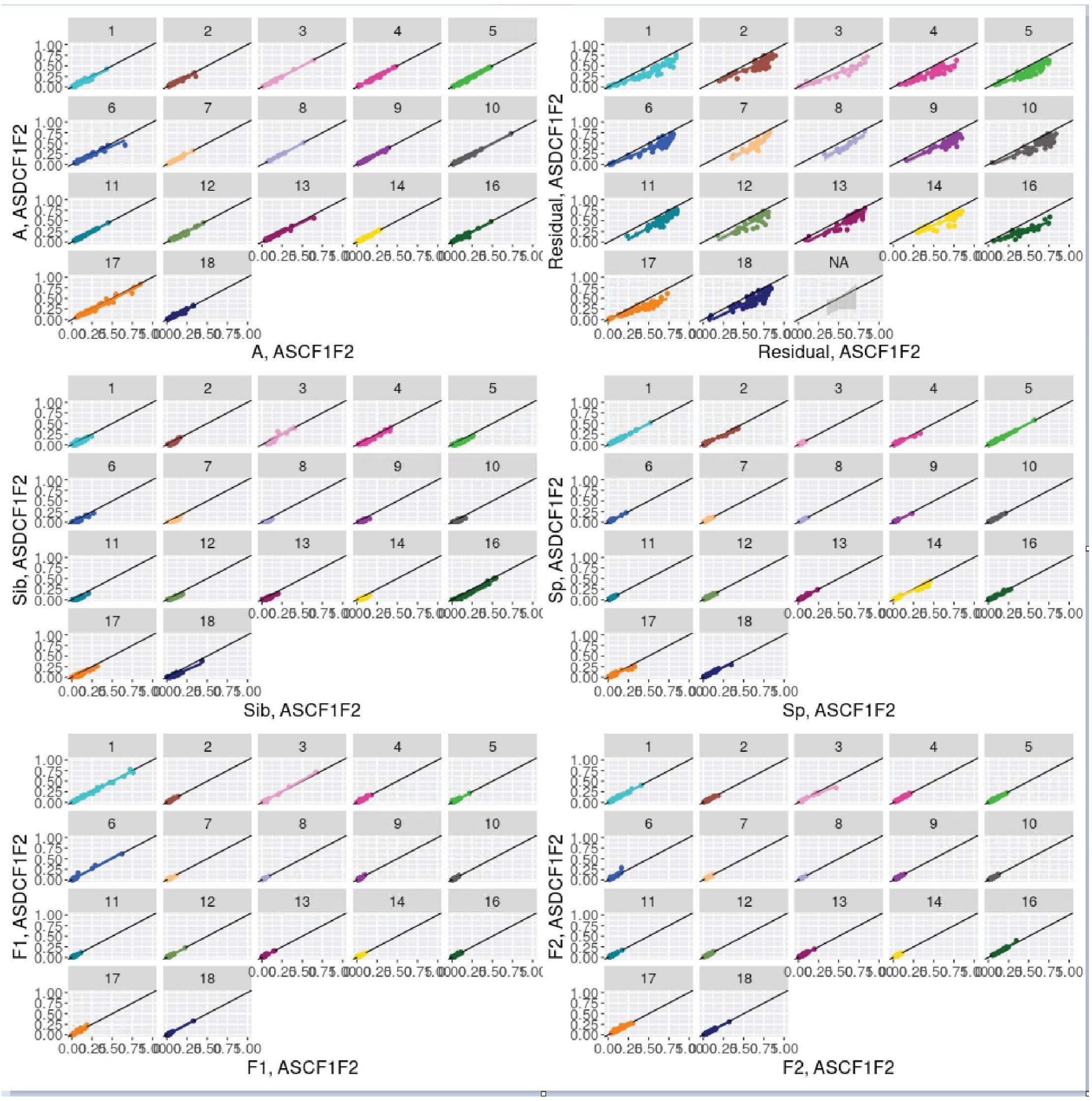


Supplementary Figure 8: Source of variance absorbed into dominance component.

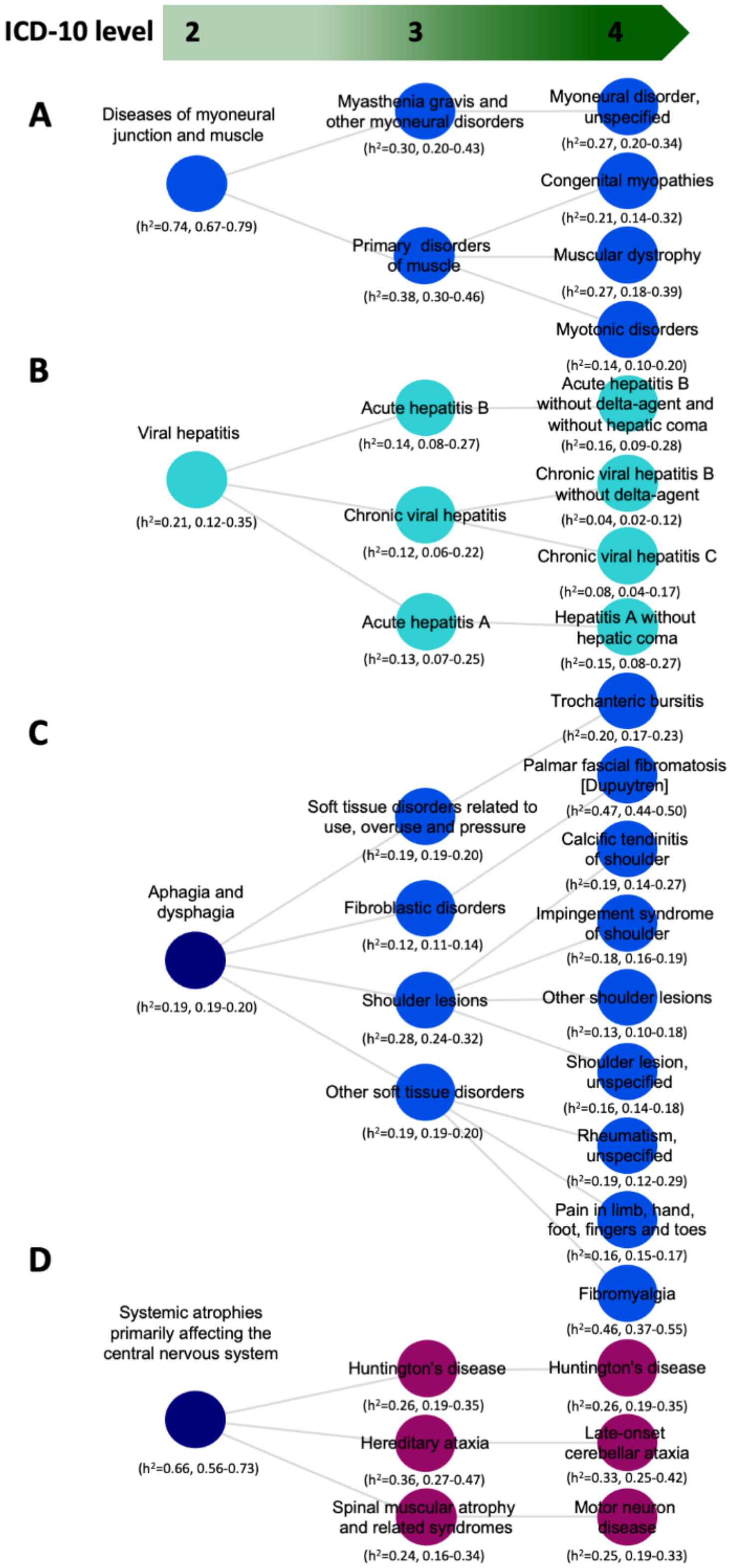


Supplementary Figure 9: Other examples from ICD where heritability increases as one goes up

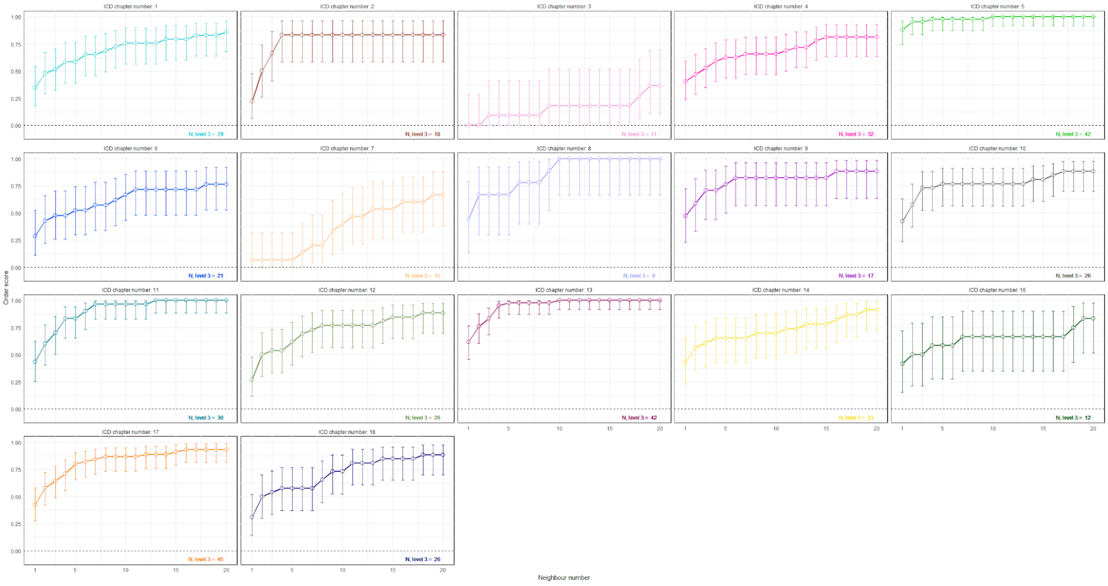


Supplementary Figure 10: Disorder score with increasing number of neighbors

#### Tables

Table 1: Comparison of heritabilities based on previous studies

| Study | Country | Population | #Phenotypes | Design | A | ASCF |
|---|---|---|---|---|---|---|
| | | | | | Pearson correlation | |
| Wang et al[4] | USA | ~481k | 124 | Nuclear Family | 0.47 (0.33-0.60) | 0.35 (0.19-0.50) |
| Mucci et al[7] | Denmark, Finland, Norway, Sweden | ~203k | 35 | Twin | 0.57 (0.11-0.83) | 0.67 (0.25-0.88) |
| Polubriaginof et al[5] | USA | ~1.1M | 136 | Extended family | 0.33 (0.19-0.45) | 0.16 (0.01-0.31) |

#### Appendix

Let $S_1, S_2 \sim \mathcal{N}(0,1)$. Assume that $\ell_1 = S_1\sqrt{1-c^2} + cS_2$ and $\ell_2 = S_2$. Then $V\ell_1 = V\ell_2 = 1$ and $\mathrm{cov}(\ell_1, \ell_2) = c$.

Let $Y_1 = 1(\ell_1 + f_1 \geq 0)$ and $Y_2 = 1(\ell_2 + f_1 \geq 0)$

$$\begin{aligned} E(Y_1Y_2) &= P(Y_1 = Y_2 = 1) = P(S_1\sqrt{1-c^2} + cS_2 + f_1 \geq 0, S_2 + f_2 \geq 0) \\ &= \int_{-f_2}^{\infty}\int_{-\frac{f_1+cs_2}{\sqrt{1-c^2}}}^{\infty} \phi(s_1)\phi(s_2)ds_1ds_2 \\ &= \int_0^1\int_0^1 \phi\left(-f_2 + \frac{x_2}{1-x_2}\right)\phi\left(-\frac{f_1 + c(-f_2+\frac{x_2}{1-x_2})}{\sqrt{1-c^2}} + \frac{x_1}{1-x_1}\right)\frac{1}{(1-x_1)^2}\frac{1}{(1-x_2)^2}dx_1dx_2 \end{aligned}$$

where we use the fact that

$$\int_a^{\infty} f(x)dx = \int_0^1 f\left(a + \frac{t}{1-t}\right)\frac{1}{(1-t)^2}dt$$

We have that $c = \sum_{i=1}^d \frac{p_ic_i}{\sum_{j=1}^d p_j}$. So

$$\begin{aligned} &\frac{\partial}{\partial p_i}E(Y_1Y_2) \\ &= \frac{\sum_{j=1}^d (p_jc_i - p_jc_j)}{\left(\sum_{k=1}^d p_k\right)^2}\int_0^1\int_0^1\left(\frac{\partial}{\partial c} - \frac{f_1 + c\left(-f_2 + \frac{x_2}{1-x_2}\right)}{\sqrt{1-c^2}}\right)\phi\left(-f_2 + \frac{x_2}{1-x_2}\right) \\ &\phi'\left(-\frac{f_1 + c(-f_2+\frac{x_2}{1-x_2})}{\sqrt{1-c^2}} + \frac{x_1}{1-x_1}\right)\frac{1}{(1-x_1)^2}\frac{1}{(1-x_2)^2}dx_1dx_2 \\ &= \frac{\sum_{j=1}^d (p_jc_i - p_jc_j)}{\left(\sum_{k=1}^d p_k\right)^2}\int_0^1\int_0^1 -\frac{f_1c(x_2-1) - f_2(x_2-1) - x_2}{(1-c^2)^{3/2}(x_2-1)}\phi\left(-f_2 + \frac{x_2}{1-x_2}\right) \\ &\phi'\left(-\frac{f_1 + c(-f_2+\frac{x_2}{1-x_2})}{\sqrt{1-c^2}} + \frac{x_1}{1-x_1}\right)\frac{1}{(1-x_1)^2}\frac{1}{(1-x_2)^2}dx_1dx_2 \end{aligned}$$

and

$$\frac{\partial}{\partial p_i}\mathrm{Loss}_\theta = 2\sum_{k>r}\left(E(Y_kY_r) - Y_kY_r\right)\frac{\partial}{\partial p_i}E(Y_kY_r)$$
